# Combining seasonal malaria chemoprevention with novel therapeutics for malaria prevention: a mathematical modelling study

**DOI:** 10.1101/2025.01.06.24319074

**Authors:** Lydia Braunack-Mayer, Josephine Malinga, Narimane Nekkab, Sherrie L Kelly, Jörg J Möhrle, Melissa A Penny

## Abstract

Vaccines, monoclonal antibodies, and long-acting injectables are being developed to prevent *Plasmodium falciparum* malaria. These therapeutics may target multiple stages of the parasite life cycle; evidence is needed to articulate their benefits with chemoprevention and prioritise candidates for clinical development. We used an individual-based malaria transmission model to estimate the health impact of combining new therapeutics with seasonal malaria chemoprevention (SMC). Our modelling framework used emulator-based methods with models of pre-liver and blood stage therapeutic dynamics. We evaluated the benefit of combining therapeutics with SMC in children under five by estimating reductions in the cumulative incidence of uncomplicated and severe malaria, relative to SMC or the new therapeutic alone, during and five years after deployment. New therapeutics may require extended pre-liver stage duration or multi-stage activity to combine with SMC. For three SMC cycles in a high transmission setting, a pre-liver stage therapeutic with partial initial efficacy (>50%) required a protection half-life >230 days to reduce cumulative severe cases by >5% five years after deployment stopped (>23% during interventions). Longer protection was needed when combined with four or five SMC cycles. Combining SMC with a multi-stage therapeutic increased public health impact both during and after deployment. Combining SMC with malaria therapeutics active against multiple stages of the parasite life cycle can improve the effectiveness of SMC, highlighting the need to prioritise the clinical development of these therapeutics for combination with malaria chemoprevention.

## Introduction

The first five years of a child’s life play a pivotal role in shaping their long-term health and development, and early childhood illness can significantly impact a child’s ability to learn and succeed later in life [1]. Nevertheless, in regions of the world where *Plasmodium falciparum* malaria is endemic, this crucial period in a child’s life coincides with their highest vulnerability to malaria infection. The World Health Organization estimates that, in 2022, approximately 452 000 children under the age of five years died of malaria in Africa [2]. In Sub-Saharan regions of Africa, public health interventions, such as insecticide treated nets [3] and chemoprevention [4], are crucial to provide children with tools that prevent malaria during these formative years.

In a number of African regions, the high-risk period for *Plasmodium falciparum* malaria transmission coincides with the rainy season. The World Health Organization recommends seasonal malaria chemoprevention (SMC), which clears existing infections and prevents new cases through monthly cycles of an antimalarial drug [4]. The drug combination sulfadoxine-pyrimethamine with amodiaquine (SP-AQ) has been widely used for SMC in children [4], achieving a 88.2% (95% CI 78.7–93.4) mean reduction in the incidence of clinical malaria in children younger than five years when deployed at scale [5]. However, children do not always comply with the full treatment course of SP-AQ, high levels of coverage are not always achieved, and SMC’s effectiveness may be compromised due to the spread of partial *Plasmodium falciparum* resistance to SP [5,6]. These gaps leave some children at risk of malaria infection during a crucial period in their development.

In 2021, the first randomised control trial was completed for SMC with SP-AQ in combination with malaria vaccination [7]. This study combined up to five cycles of an established chemoprevention intervention with seasonal deployment of the first approved malaria vaccine, RTS,S/AS01E (RTS,S). SP-AQ acts to clear blood stage parasites and prevent liver stages from emerging into the blood, while RTS,S is a pre-liver stage vaccine that prevents liver stage infection by targeting surface antigens of sporozoite parasites. The combination of a pre-liver stage vaccine with SMC was found to be superior to SMC alone [7–9]. SMC has also been studied in combination with the R21/Matrix-M malaria pre-liver stage vaccine [10]. And, modelling studies have identified the potential for substantial public health benefits from combining RTS,S with SMC [11]. These successes demonstrate that a pre-liver stage vaccine can be used to strengthen the effectiveness of malaria chemoprevention and to achieve significant reductions in childhood burden. However, it is not yet known whether alternative therapeutics will provide similar benefits, or whether therapeutics that target additional stages of the parasite life cycle could be more effective.

Recent developments in vaccines, monoclonal antibodies, and long-acting injectable drugs suggest opportunities to combine chemoprevention with therapeutics other than pre-liver stage vaccines, including with therapeutics active against multiple stages of the parasite life cycle. The first phase two clinical trial results for a pre-liver stage monoclonal antibody are now available [12]. This class of therapeutic can provide high levels of protection following a single dose, making it a likely candidate for seasonal deployment. But concerns remain regarding the cost-effectiveness of monoclonal antibodies when deployed at scale [13,14]. Injectable formulations of existing antimalarial drugs, such as atovaquone-proguanil, may offer children three to four months of protection at a lower cost than monoclonal antibodies [14–17]. These re-formulations can target multiple stages of the parasite life cycle by, for example, combining drug partners with both liver stage and blood stage activity. There are also candidates for multi-stage in development, including whole-sporozoite [18], blood stage [19], and transmission blocking vaccines [20]. Each of these novel therapeutics could be combined with SMC to improve the effectiveness of childhood malaria interventions. But each of these novel therapeutics has different profiles, encompassing differences in the levels of protection provided immediately following administration, the length of protection afforded, and their rate of efficacy decay. It is not yet known how these differences in therapeutic properties will relate to their public health impact once combined with SMC.

In this modelling study, we explored and defined the essential properties needed for a seasonally deployed, novel malaria therapeutic to increase the effectiveness of SMC throughout childhood. By novel therapeutic, we refer to a malaria vaccine, monoclonal antibody, or long-acting injectable drug that has not yet been approved for use in children. Our research combined an established, individual-based malaria transmission model with statistical modelling techniques, quantifying the therapeutic properties required for a new therapeutic to contribute additional reductions in cumulative uncomplicated and severe cases in children, in comparison to the standard of care. Our results encompassed vaccines, monoclonal antibodies, and novel long-acting therapeutics with both activity against a single-stage of the parasite life cycle (either pre-liver or blood stage) and activity against multiple stages (pre-liver combined with blood stage, referred to as multi-stage therapeutics). Through this evidence, we have aimed to provide funders, policymakers, and developers with guidance to inform candidate selection of malaria therapeutics with adequate properties to meet public health goals in advance of phase two and three clinical trials.

## Materials and Methods

### Malaria transmission model

We used an established model of malaria transmission to predict the impact of combining SMC with seasonal deployment of a novel therapeutic. This individual-based model, called OpenMalaria (https://github.com/SwissTPH/openmalaria/wiki), has been fully described previously [21]. OpenMalaria combines multiple, smaller models that each represent a component in the malaria transmission lifecycle. Model components simulate the dynamics of mosquito populations, malaria parasitaemia in the course of an infection, onward transmission, and the processes leading to clinical illness, severe disease, and death (Supplement Table 1.1). This model has been validated to historical data on a range of epidemiological outcomes, including patterns of age-specific prevalence and incidence, and age-specific rates for malaria mortality and hospitalisation [21].

Importantly for this study, OpenMalaria includes detailed within-host models that represent parasite dynamics across multiple stages of the *Plasmodium falciparum* life cycle. The model simulates the probability of liver stage infection following an infectious mosquito bite, the resulting blood stage parasite density, and the probability of onward transmission. Therapeutics can target each of these stages independently. As a result, synergistic effects of combining interventions on population-level outcomes, such as those observed in the clinical trial of RTS,S/AS01 and SMC,[7–9] can arise from the underlying model mechanisms, rather than being imposed at an individual level through assumed interaction terms or fixed efficacy profiles. This allows interactions between therapeutics, including vaccines and drugs, to emerge at the population level as a consequence of their combined effects on parasite development and host immunity.

Finally, OpenMalaria’s model components capture the dynamics of acquired immunity to the asexual blood stages of *Plasmodium falciparum* infection. Acquired immunity is represented as a function of an individual’s cumulative density of asexual parasitaemia since birth, including maternal immunity, and their total number of prior infections. Infection densities are drawn from parasitaemia records of malaria-naive patients infected with *Plasmodium falciparum* for treatment of neurosyphilis [22–24]. The number of prior infections influences pre-erythrocytic immunity, and both prior infections and cumulative asexual parasitaemia influence blood stage immunity. Immunity acts by reducing an infected host’s parasite densities and affecting the probability that parasitaemia will reach the specific parasite densities, or pyrogenic thresholds, that trigger symptoms of uncomplicated and severe malaria [23,24].

### Intervention dynamics and deployment

To model SMC with SP-AQ, we represented the drug combination with both preventative and curative effects. SP-AQ was assumed to prevent new infections by blocking liver-stage parasite emergence and to cure existing infections by clearing blood and liver-stage parasites at the time of administration. These assumptions followed our review of the mechanisms of action of SP-AQ for malaria prevention [25]. Efficacy decay for the preventative action was calculated with a Weibull function: E(t) = E(0) × exp(−(t/L)^k^ × log(2)), where E(0) was the initial probability of preventing infection at the time of administration, L the scale parameter was the number of days until this probability reached half of its initial value, and k the shape parameter influenced the rate of efficacy decay.

To model novel malaria prevention therapeutics, we simulated a range of malaria vaccine, monoclonal antibody, and long-acting injectable drug candidates by combining explicit models of therapeutic activity against different stages of the *Plasmodium falciparum* parasite life cycle. Pre-liver stage therapeutics were assumed to reduce the probability of infection following exposure, while blood stage therapeutics killed circulating parasites. Multi-stage therapeutics were modelled by combining both components. As for SP-AQ, the efficacy of each component was represented with a Weibull function of time following administration. These model parameters are illustrated in supplement Fig 1.1.

For each therapeutic (SP-AQ, pre-liver stage, blood stage, and multi-stage profiles), Table 1 outlines the modelled range of parameters used. For SMC, model parameter values were previously calibrated to data from a randomised non-inferiority trial of SMC with SP-AQ [26,27]. For novel therapeutics, we sampled 500 sets of parameter values using Latin hypercube sampling with a uniform probability distribution. Parameter ranges were chosen to represent the broadest possible range of therapeutic profiles, from a highly efficacious, long-acting vaccine (initial efficacy 100%, protection half-life 500 days, exponential decay shape of 1) to a low-efficacy, short-acting drug (30% initial efficacy, 30 day protection half-life, rapid decay shape of 9). By simulating outcomes for this broad parameter space of protective properties, we were able to explore the impact of combining SMC with a full spectrum of possible therapeutics. This space aimed to encompass the properties of future novel vaccines, monoclonal antibodies, and long-acting injectable drugs.

**Table 1.**
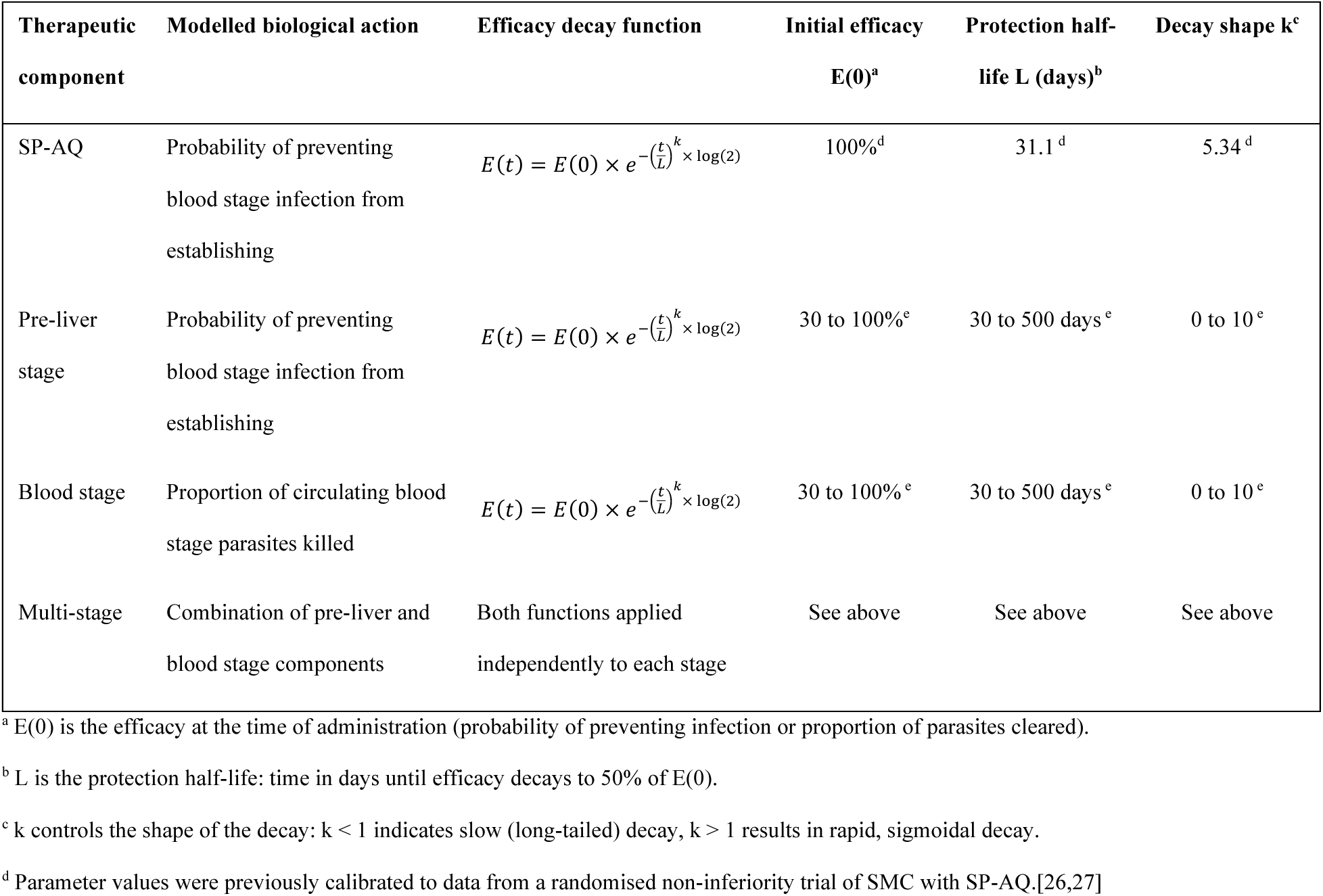
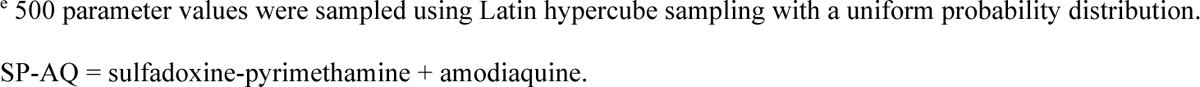
Model assumptions for dynamics of therapeutic efficacy and decay.

We then used our individual-based model of malaria transmission to simulate the deployment of SMC with SP-AQ, both alone and in combination with novel malaria prevention therapeutics. The novel therapeutic (pre-liver stage, blood stage, multi-stage) was deployed together with the distribution of SP-AQ in the first cycle of SMC, representing a scenario where a child receives a vaccine, monoclonal antibody or long-acting injectable in combination with receipt of SP-AQ. All interventions were deployed to children aged between three and 59 months of age. Importantly, as described above, we did not make any assumptions about drug-drug or vaccine-vaccine interactions, synergisms or antagonisms between the two components of multi-stage therapeutics; any synergistic effects on clinical outcomes at population-level emerged from the underlying within-host intervention dynamics of OpenMalaria.

### Intervention model validation

In a previous study, we validated our model’s ability to accurately represent the effectiveness of SMC in combination with a pre-liver stage vaccine [28]. We used OpenMalaria to replicate outcomes from the only available clinical trial evaluating this combination: the individually randomised trial of RTS,S with or without SMC conducted in Mali and Burkina Faso [7,8]. Using data from the first two years of the trial, we calibrated vaccine booster efficacy and the therapeutic properties of SP-AQ to match observed clinical incidence in all three trial arms: SMC alone, RTS,S alone, and the combination. This was done with a Bayesian optimisation framework within OpenMalaria, taking initial efficacy estimates for RTS,S after the three-dose primary series (91.1%) and a half-life of 7.3 months from previous modelling studies and clinical trial data [29]. These values were used as inputs, and boosting efficacies were estimated by optimising model predictions against trial data.

OpenMalaria successfully reproduced monthly clinical incidence and hazard ratios for all arms of the trial in both countries. For RTS,S booster doses, the model estimated that a 12-month post-primary booster restored efficacy to 78.96% (95% CI 70.74 – 87.18%) in the vaccine-only arm and 77.45% (95% CI 68.26 – 86.62%) in the combination arm. A second booster reduced maximum efficacy to 57.31% (95% CI 44.05 – 70.56%) and 48.76% (95% CI 34.57 – 62.95%), in the vaccine only and combination arms respectively. These values matched closely with observed outcomes, and our model captured both temporal trends and relative effectiveness across trial arms.

### Outcome measures

Our primary outcome measures captured reductions in the cumulative incidence of uncomplicated and severe malaria throughout childhood. We followed children born in the first year of intervention delivery and evaluated cumulative incidence at five (as the intervention ends) and ten years old (five years after interventions have ended). As illustrated in Fig 1, we then calculated the reductions in the cumulative incidence of uncomplicated and severe malaria attributed to the combination of SMC with SP-AQ and the novel therapeutic. Reductions were evaluated relative to two counterfactual scenarios: regular deployment of SMC alone, and seasonal deployment of a novel therapeutic alone (Supplement Section 1). We report these reductions as the percentage of burden averted by the combination relative to a standard of care; we do not compare to a no-intervention scenario.

**Fig 1.**
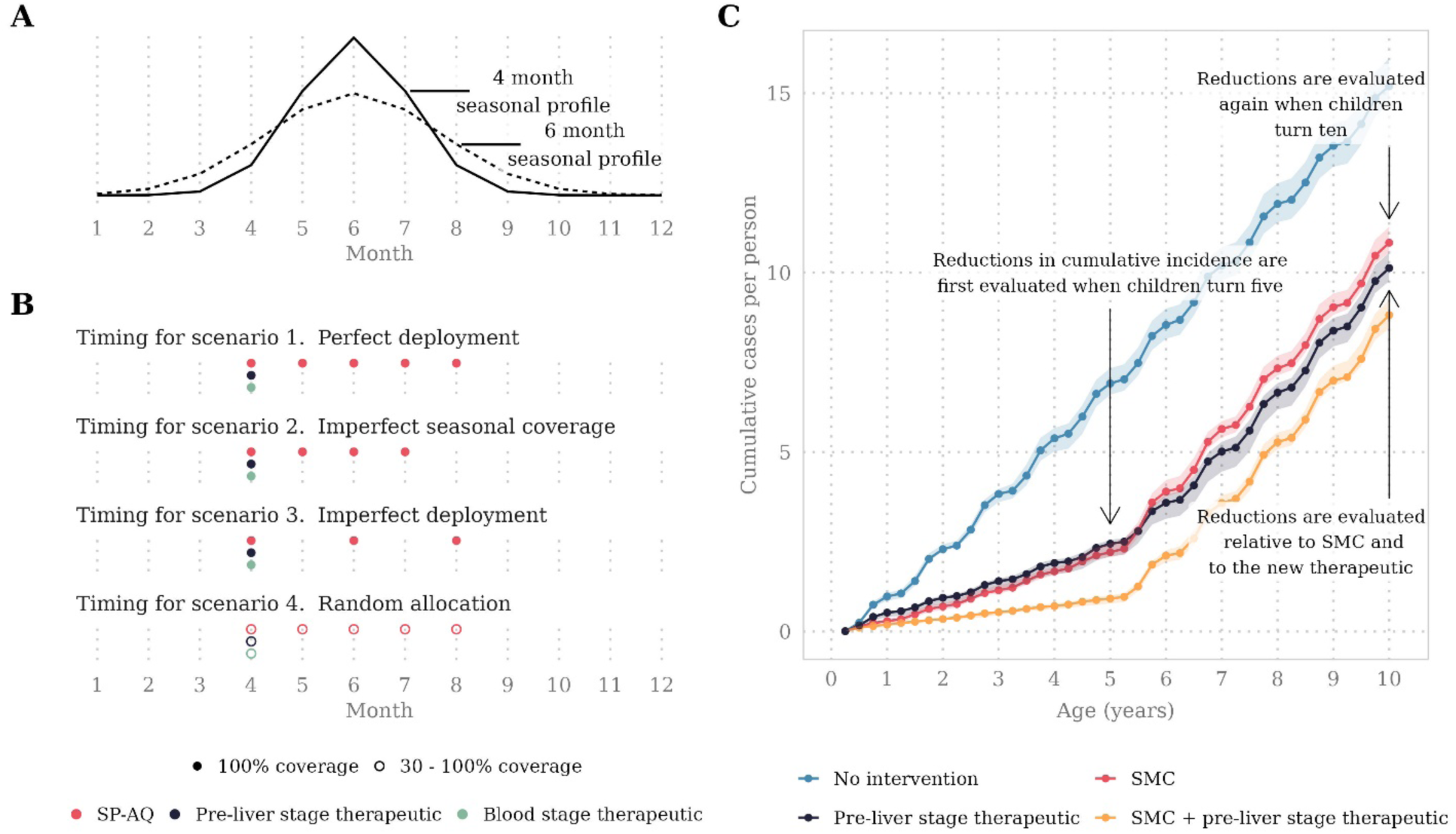
Illustration of seasonality profiles, intervention timing, and primary outcome measures. Panel A illustrates the two seasonal profiles modelled, where 70% of malaria cases occur within a four- or six-month period and where the y-axis represents the average annual entomological inoculation rate (EIR). Panel B illustrates interventions deployed according to four seasonal deployment scenarios. Scenario 1. Perfect deployment: All children aged between three and 59 months had perfect compliance to five yearly SMC cycles. Scenario 2. Imperfect seasonal coverage: All children aged three to 59 months had perfect compliance to four yearly SMC cycles but missed the final cycle. Scenario 3. Imperfect deployment: All children aged three to 59 months had perfect compliance to yearly SMC cycles one, three, and five. Scenario 4. Random allocation: 30% to 100% of children were randomly selected to receive each of five yearly SMC cycles. Panel C illustrates the evaluation of our primary outcome measures. We tracked a cohort of children born in the first year of intervention deployment, and evaluated cumulative incidence when children were five and ten years old. We then calculated the reductions in the cumulative incidence of uncomplicated and severe malaria attributed to the combination of SMC and the novel therapeutic. Reductions were evaluated relative to two counterfactual scenarios: regular deployment of SMC alone, and seasonal deployment of a novel therapeutic alone. Shaded regions indicate the minimum and maximum cumulative cases per person observed across stochastic replicates of simulations from the individual-based malaria transmission model.

### Simulation and model scenarios

The malaria transmission model was used to simulate cumulative case outcomes for a range of transmission and health system scenarios. Each model scenario captured a unique combination of the following characteristics: a low (10%) or high (50%) probability of seeking first-line treatment for clinical malaria over 14 days post-infection; 70% of malaria cases occurring within a four- or six-month transmission season (Fig 1); six levels of transmission intensity ranging from low-moderate (9%) to high (64%) baseline annual prevalence. Annual prevalence was measured as the prevalence rate of *Plasmodium falciparum* infections detectable by rapid diagnostic test in children aged two to ten years old (*Pf*PR_2-10_) when no intervention was deployed.

We also modelled four different seasonal deployment scenarios for SMC, as illustrated in Fig 1. In the perfect deployment scenario, all children aged between three and 59 months had perfect compliance to five yearly SMC cycles. This scenario represented an ideal clinical trial where every child is perfectly adherent to the SP-AQ chemoprevention regimen. In the imperfect seasonal coverage scenario, all children aged three to 59 months had perfect compliance to four yearly SMC cycles but missed the final cycle. This scenario represented a setting where the number of SMC cycles may not provide protection for the full transmission season. In the imperfect deployment scenario, all children aged three to 59 months had perfect compliance to yearly SMC cycles one, three, and five.

This scenario represented a setting where children systematically miss out on multiple SMC cycles. In the random allocation scenario, 30% to 100% of children were randomly selected to receive each of five yearly SMC cycles. This scenario represented a setting where children randomly miss out on one or more SMC cycles. In all four deployment scenarios, the novel prevention intervention was co-deployed seasonally together with the first cycle of SMC. By varying deployment, we were able to quantify differences in the novel intervention’s benefit when children are more or less covered by SMC.

### Statistical analysis

We followed an existing mathematical framework for defining essential therapeutic characteristics for malaria prevention interventions [30]. This framework has previously been used to explore determinants of malaria intervention impact for long-acting injectable drugs [26] and for novel seasonal malaria chemoprevention therapeutics [31].

In brief, following our methodology (Supplement Section 1, Fig 1.2), we first generated a Latin hypercube sample of 500 values for the novel intervention’s parameters: therapeutic initial efficacy, protection half-life, and decay shape. For each sample, we used our individual-based malaria transmission model to simulate outcomes with ten stochastic replicates. Heteroskedastic Gaussian process regression models [32] were then fitted to predict primary outcome measures (reductions in cumulative uncomplicated and severe cases at five and ten years old) as a function of intervention inputs (initial efficacy, protection half-life, and decay shape). These models were used to emulate the relationship between therapeutic properties and our primary outcome measures without additional simulation from the individual-based model, which would have required intensive computation. Emulator performance was evaluated against a 10% hold-out set. Finally, for each model scenario, we quantified the impact of a small change in each key therapeutic property on variation in our primary outcome measures by performing a non-parametric sensitivity analysis using the Sobol-Jansen method [33,34]. Sobol total-order indices were computed for two Latin hypercube samples of 50 000 intervention parameters. All analyses were conducted in R [35].

## Results

### Combining SMC with seasonal deployment of a pre-liver stage therapeutic

We first simulated cumulative case outcomes among children for exemplar malaria therapeutic profiles: one pre-liver stage therapeutic with long duration and high efficacy (protection half-life 354 days, 90% initial efficacy) and a second with shorter duration and partial efficacy (protection half-life 120 days, 50% initial efficacy). This allowed us to explore the therapeutic properties needed for a pre-liver stage therapeutic to add benefit to SMC with SP-AQ. Fig 2 depicts the cumulative incidence curves predicted by our model for two hypothetical pre-liver stage therapeutics. First, we saw that both therapeutics contributed to reductions in cumulative uncomplicated and severe cases by the time children were five years old (reductions >10%). This result confirmed the positive effects observed in the randomised control trial of seasonal vaccination with RTS,S in combination with SMC,[8,9] and the R21 phase three clinical trial.[10]

**Fig 2.**
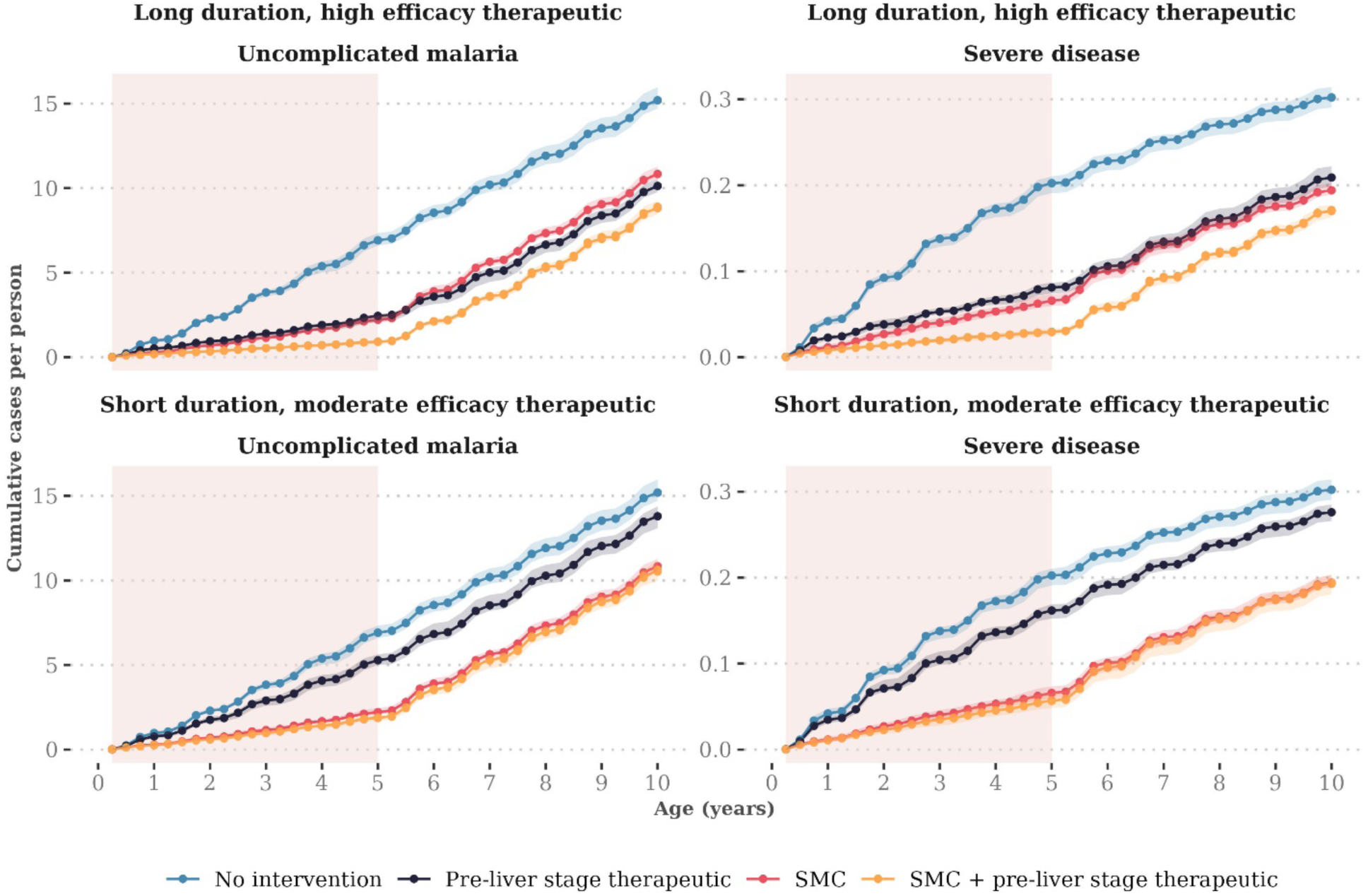
Predicted cumulative case curves when seasonal deployment of two exemplar pre-liver stage therapeutics are combined with SMC. The median number of cumulative cases per year of age were calculated when children aged three to 59 months received one of the following interventions: no intervention, three SMC cycles (imperfect deployment scenario), seasonal deployment of a pre-liver stage therapeutic, or the combination of three SMC cycles and the pre-liver stage therapeutic. Top panels include deployment of a pre-liver stage therapeutic with a protection half-life of 354 days, 90% initial efficacy, and decay shape parameter of 1. Bottom panels include a pre-liver stage therapeutic with a protection half-life of 120 days, 50% initial efficacy, and decay shape parameter of 1. The pink shaded regions indicate age-eligibility for SMC and the novel therapeutic. Shaded regions indicate the minimum and maximum cumulative cases per person observed across stochastic replicates of simulations from the individual-based malaria transmission model. Model results are shown for a high transmission scenario (32% *Pf*PR2-10) where 75% of malaria cases occur within six months of the year and the probability of seeking first-line treatment for clinical malaria over 14 days is low (10%).

However, we saw that pre-liver stage therapeutics needed to have a long duration and high efficacy to maintain their benefits in comparison to SMC as children aged. The exemplar therapeutic with shorter duration and partial efficacy (Fig 2) led to no observable difference in cumulative uncomplicated cases by the time children were eight years old.

Furthermore, we observed that the combination of SMC with SP-AQ and the pre-liver stage therapeutic was more beneficial than the pre-liver stage therapeutic alone throughout childhood. Similar patterns were observed for other SMC deployment scenarios (Supplement Fig 2.1-2) and across the full range of pre-liver stage therapeutic properties (Supplement Fig 2.3). Together, these findings indicate that pre-liver stage therapeutics may need a long duration to limit the potential for delayed malaria when added to SMC and may not fully replace SMC’s benefits.

In addition to a long duration, we found that pre-liver stage therapeutics needed a long tail of protection to maintain reductions in cumulative burden as children aged. Higher reductions in cumulative cases by ten years of age were observed for pre-liver stage therapeutics with a long tail of protection, encompassing exponential and biphasic decay profiles (decay shape <1, Fig 3), in comparison to sigmoidal decay (decay shape >1). For example, in Fig 3, deploying a pre-liver stage therapeutic with a decay shape parameter between 0.6 and 0.8 was associated with a 9.9% median reduction (interquartile range 5.5-16.0% across the parameter range for protection half-life and initial efficacy) in cumulative uncomplicated cases by the time children were ten years old, evaluated relative to SMC alone. In contrast, deploying a therapeutic with a decay shape between 4.0 and 4.2 led to a lower median reduction of 5.3% (interquartile range 1.8-10.0%). Similar findings were observed across deployment scenarios (Supplement Fig 2.4-2.6).

**Fig 3.**
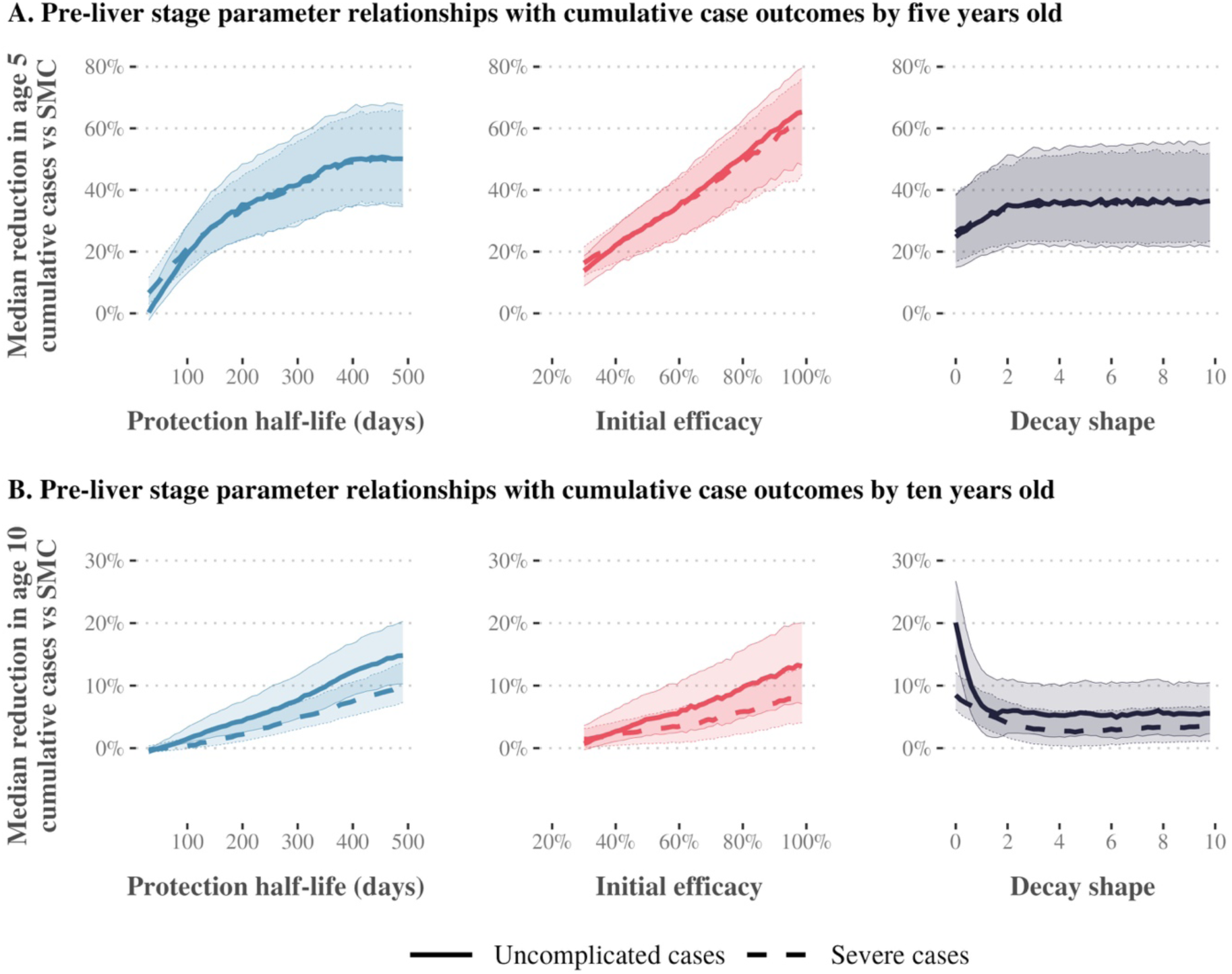
Gaussian Process regression emulator predictions for the relationship between pre-liver stage therapeutic properties and reductions in cumulative uncomplicated and severe cases relative to SMC alone. Results show the imperfect deployment scenario, where children aged three to 59 months received three SMC cycles. Transmission is high (32% *Pf*PR2-10), 75% of malaria cases occur within six months of the year, and the probability of seeking first-line treatment for clinical malaria over 14 days is low (10%). Each panel shows the median reduction in cumulative uncomplicated (solid lines) or severe cases (dashed lines) achieved by combining seasonal deployment of a pre-liver stage therapeutic with SMC, relative to cumulative cases when SMC is deployed alone. Median reductions are calculated by dividing the parameter range for the therapeutic property shown on the x-axis (protection half-life, initial efficacy, decay shape) into 51 segments, and calculating the median outcome for each given segment across all other parameter values. Shaded regions represent the 25th and 75th percentiles of the corresponding reductions. Predictions are shown separately for cumulative case outcomes at five (top panels) and ten years old (bottom panels). SMC = seasonal malaria chemoprevention..

Our results showed that co-deploying SMC with a pre-liver stage therapeutic is likely to have the most impact in settings where SMC does not fully protect children. For example, for a pre-liver stage therapeutic with 50% initial efficacy and a long tail of protection (biphasic decay shape 0.7) deployed with three yearly cycles of SMC, a target 5% reduction in cumulative severe cases by ten years old required a protection half-life of at least 237 days (Fig 4). For the same therapeutic profile, a protection half-life of at least 237 days corresponded to a reduction in severe cases by five years old of more than 23%. When the same therapeutic was combined with five SMC cycles, however, a 5% reduction required a protection half-life of at least 340 days. Results also indicated that higher initial efficacy could be traded for shorter duration requirements; in Fig 4, the target 5% reduction in cumulative severe cases by ten years old could be achieved with a protection half-life of 153 days if pre-liver stage initial efficacy was 70%. Similar results were observed across seasonal transmission settings and levels of access to care. (Supplement Fig 2.7-8)

**Fig 4.**
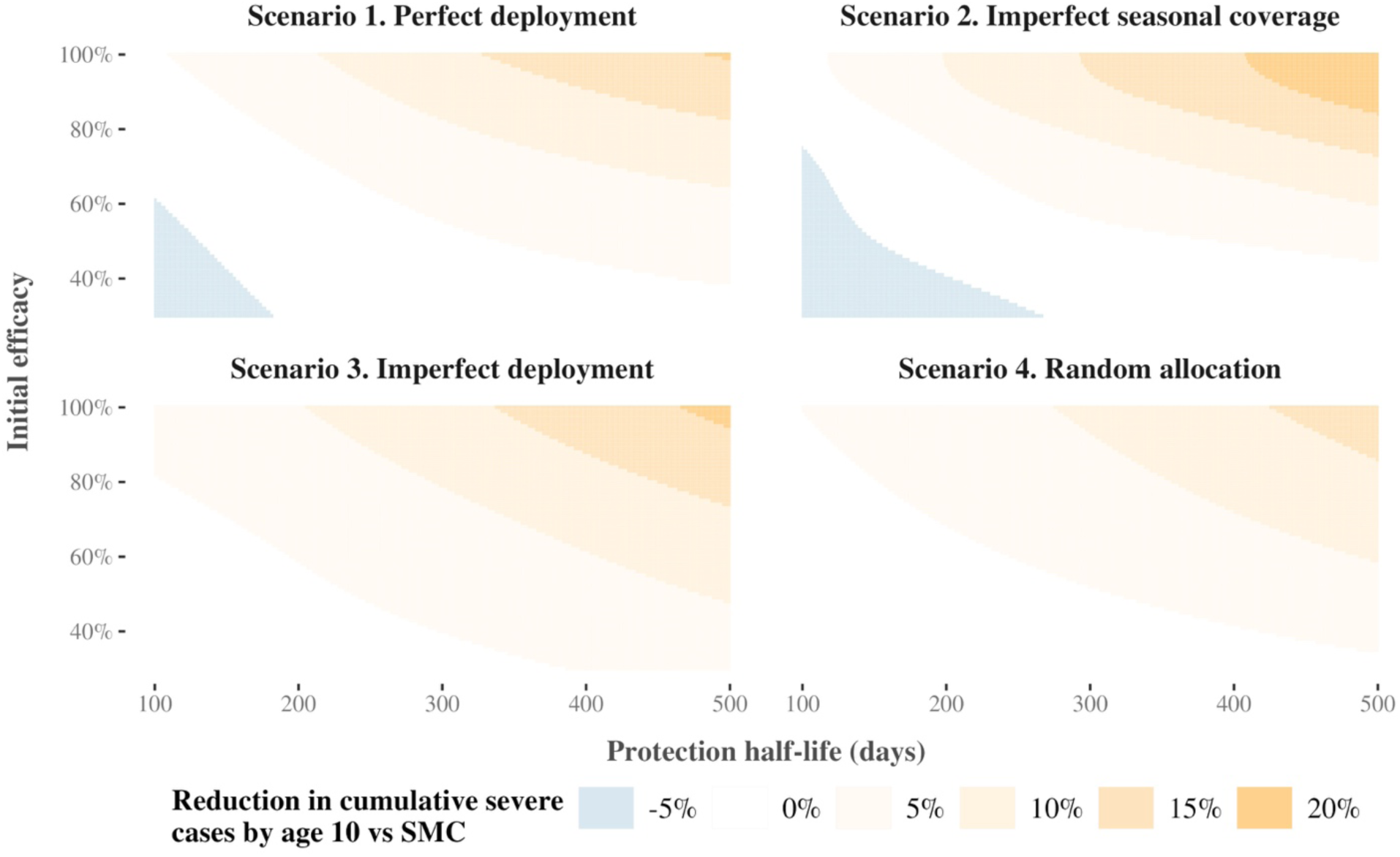
Predicted relationships between pre-liver stage therapeutic properties and reductions in cumulative severe cases relative to SMC alone, compared across SMC deployment scenarios. Grid squares indicate the predicted reduction if a pre-liver stage therapeutic with the given protection half-life (x-axis) and initial efficacy (y-axis) were deployed in addition to SMC, assuming a long tail of protection (decay shape parameter 0.7). Reductions are rounded to the nearest 5%. Results show a scenario where transmission is high (32% *Pf*PR2-10), 75% of malaria cases occur within six months of the year, and the probability of seeking first-line treatment for clinical malaria over 14 days is low (10%). Panels depict predicted relationships for four different SMC deployment scenarios. In deployment scenario 4, SMC coverage is fixed at 70%. SMC = seasonal malaria chemoprevention, *Pf*PR2-10 = Annual *Plasmodium falciparum* parasite prevalence in children aged between 2 and 10 years old.

### Combining SMC with seasonal deployment of a blood stage therapeutic

Our results indicate that combining a blood stage therapeutic with SMC is likely to reduce the occurrence of severe malaria but may not reduce uncomplicated cases. When a blood stage therapeutic was combined with SMC, increasing a therapeutic’s initial efficacy or protection half-life led to limited change in reductions in uncomplicated malaria. As shown in Fig 5, a near-perfect initial blood stage efficacy of 95.8% was required to achieve a positive median reduction in cumulative uncomplicated cases by ten years old. In contrast, a 10% reduction in cumulative severe cases could be achieved with an initial blood stage efficacy of 86%. Similar trends were observed across SMC deployment scenarios (Supplement Fig 3.1-3.3).

**Fig 5.**
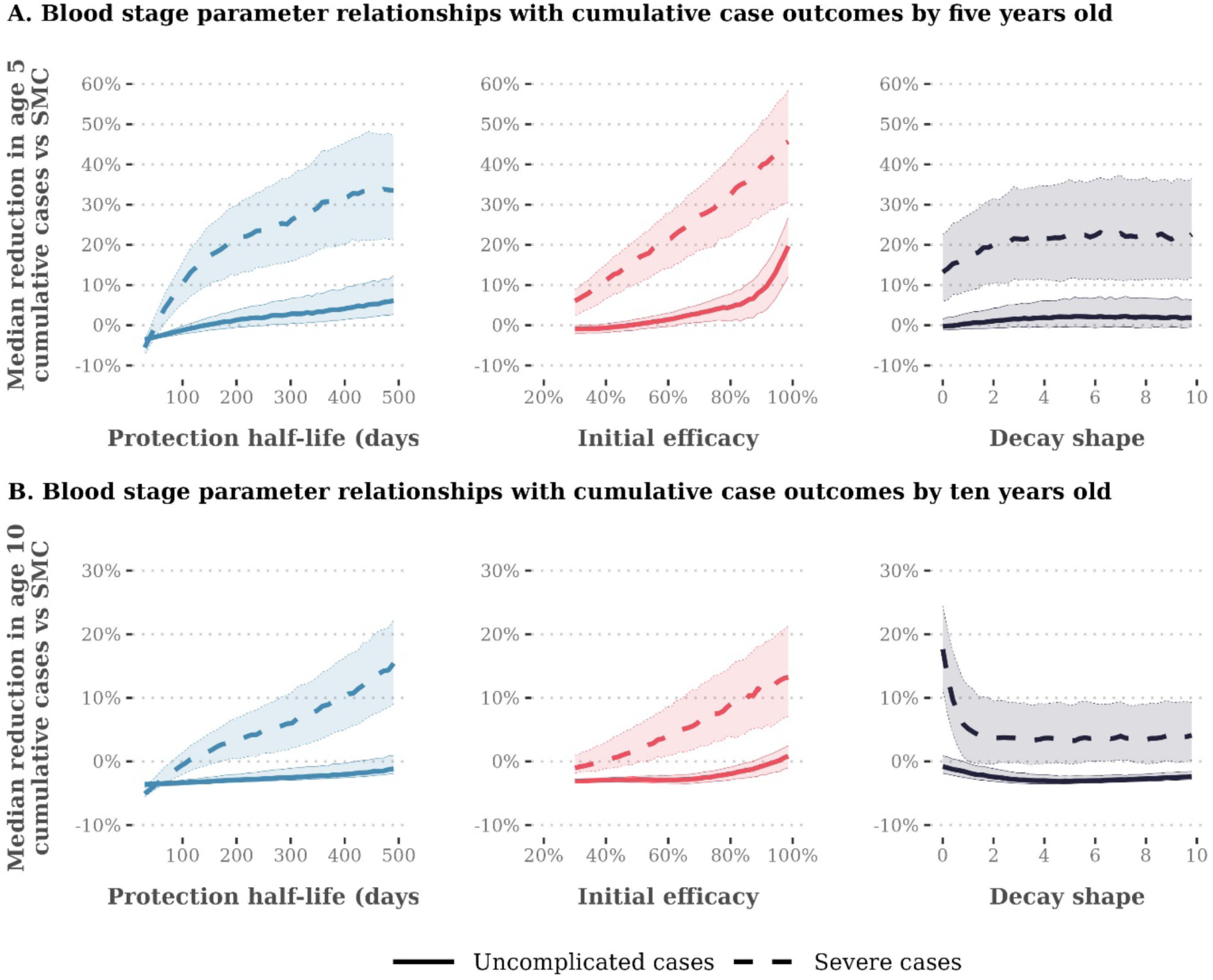
Gaussian Process regression emulator predictions for the relationship between blood stage therapeutic properties and expected reductions in cumulative uncomplicated and severe cases relative to SMC alone. Results show the imperfect deployment scenario, where children aged three to 59 months received three SMC cycles. Transmission is high (32% *Pf*PR2-10), 75% of malaria cases occur within six months of the year, and the probability of seeking first-line treatment for clinical malaria over 14 days is low (10%). Each panel shows the median reduction in cumulative uncomplicated (solid lines) or severe cases (dashed lines) achieved by combining seasonal deployment of a blood stage therapeutic with SMC, relative to cumulative cases when SMC is deployed alone. Median reductions are calculated by dividing the parameter range for the therapeutic property shown on the x-axis (protection half-life, initial efficacy, decay shape) into 51 segments, and calculating the median outcome for each given segment across all other parameter values. Shaded regions represent the 25th and 75%th percentiles of the corresponding reductions. Emulator predictions are shown separately for cumulative case outcomes at five (panels A) and ten years old (panels B). SMC = seasonal malaria chemoprevention, *Pf*PR2-10 = Annual *Plasmodium falciparum* parasite prevalence in children aged between 2 and 10 years old.

### Combining SMC with seasonal deployment of a multi-stage therapeutic

Finally, results suggest that multi-stage therapeutics with both pre-liver and blood stage activity may be preferred for interventions that aim to reduce the burden of severe malaria in children. When three SMC cycles were deployed with a multi-stage therapeutic with both pre-liver and blood stage activity, we found that pre-liver stage activity was the primary driver of impact on cumulative uncomplicated cases. More than 70% of variation in uncomplicated case outcomes was attributed to the combined impact of the pre-liver stage component’s initial efficacy and protection half-life (Fig 6, panel A). However, blood stage activity was crucial for impact on cumulative severe cases, particularly by the time children were ten years old; up to 89% of variation in the reduction in cumulative severe cases could be attributed to the blood stage component’s initial efficacy and protection half-life (Fig 6, panel A). Similar trends occurred across levels of access to care and transmission seasonality (Supplement Fig 4.1).

**Fig 6.**
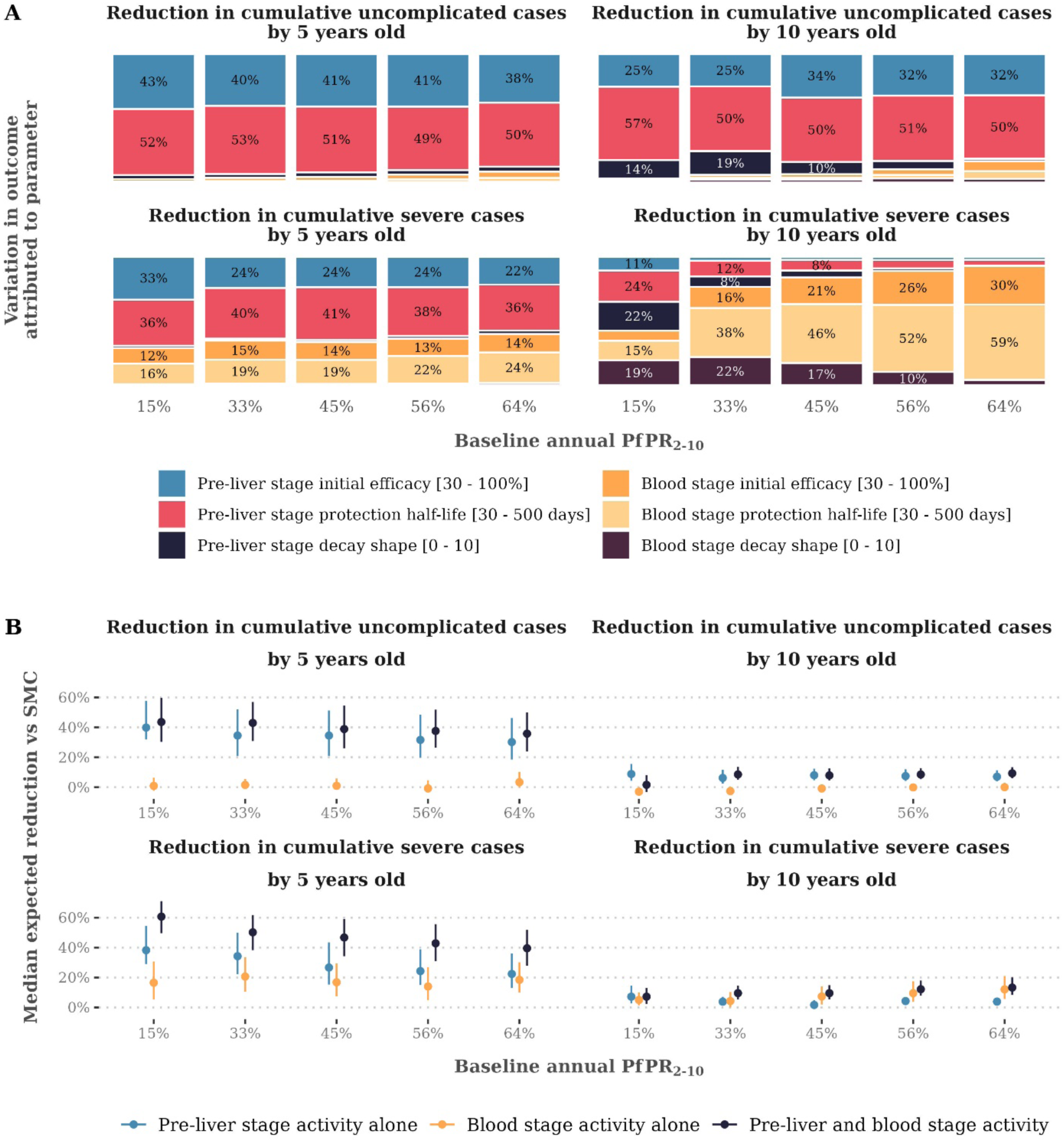
Impact of combining a therapeutic with both pre-liver stage and blood stage activity with three yearly cycles of SMC on cumulative uncomplicated and severe cases, compared with SMC deployed alone. Panel A shows the drivers of impact on all cumulative case outcomes for a therapeutic with both pre-liver stage and blood stage activity deployed together with three yearly cycles of SMC, compared with SMC deployed alone. Bars show the total Sobol effect indices for intervention model parameters. Total Sobol effect indices can be interpreted as the proportion of variation in the cumulative case outcome that can be attributed to a small change in each model parameter and its interactions with other parameters. Indices are shown across levels of transmission intensity (x-axis) for a scenario where three yearly cycles of SMC are deployed (imperfect coverage scenario) in a setting where 75% of malaria cases occur within six months of the year and the probability of seeking first-line treatment for clinical malaria over 14 days is low (10%). Panel B shows differences in the median impact on cumulative case outcomes for therapeutics with pre-liver stage activity alone, blood stage activity alone, and multi-stage pre-liver stage and blood stage activity. Results are shown for the same scenario as in panel A. Points in each panel show the median expected reduction in cumulative cases achieved by combining seasonal deployment of a pre-liver stage, blood stage, or multi-stage therapeutic with SMC, relative to cumulative cases when SMC is deployed alone. Median reductions are evaluated across all possible combinations of therapeutic initial efficacy, protection half-life, and decay shape. Bars indicate the 25th and 75th percentiles of the corresponding reductions. SMC = seasonal malaria chemoprevention, *Pf*PR2-10 = Annual *Plasmodium falciparum* parasite prevalence in children aged between 2 and 10 years old.

Importantly, model results indicated that deploying a therapeutic with combined pre-liver stage and blood stage activity could lead to greater impact than either therapeutic component alone. When children received three SMC cycles, the expected gain in median outcome between therapeutics with pre-liver stage activity alone and both pre-liver stage and blood stage activity was up to 22.5% across levels of transmission intensity for reductions by five years old, and up to 9.4% for reductions by ten years old (Fig 6, panel B). This trend occurred across SMC deployment scenarios (Supplement Fig 4.2-4.4). This result suggests that multi-stage therapeutics layered with SMC may offer substantial additional benefit for malaria outcomes in comparison to a single-stage therapeutic.

## Discussion

We used mathematical modelling to estimate the potential benefits of deploying SMC to children under five years in combination with a range of novel prevention therapeutics with both single- and multi-stage targets. We compared the benefits of the combined interventions to either SMC with SP-AQ or to the new therapeutic alone, capturing cumulative patterns in malaria uncomplicated and severe cases by five years of age, during intervention deployment, and at ten years of age, five years after deployment had stopped. To identify the therapeutic profiles that would result in added benefit from combining with SMC over SMC deployed alone, our results showed that developers of pre-liver stage therapeutics will need to prioritise candidates with a long duration (>230 days) and long tail of protection (exponential or biphasic decay). Conversely, we found that combining SMC with a new therapeutic always had added benefit compared with the therapeutic deployed alone. We also demonstrated the benefit of a therapeutic with both pre-liver stage and blood stage activity for impact on severe disease outcomes, highlighting the need for developers to prioritise multi-stage malaria therapeutics in particular.

Our results support continued investment in research and development for pre-liver stage malaria therapeutics that meet minimum duration and efficacy criteria. In particular, we found that when a pre-liver stage therapeutic was combined with three cycles of SMC in a high transmission setting, a protection half-life of more than 230 days, an initial efficacy of more than 50%, and a long tail of protection were required to maintain reductions in severe cases by ten years of age. This suggests a likely role for improved malaria vaccines in combination with SMC, rather than a pre-liver stage therapeutic with rapid decay. This work did not specifically attempt to model outcomes for either of the two approved malaria vaccines, RTS,S and R21 and we did not incorporate waning boosting efficacy into our therapeutic models. However, if the goal of deploying SMC in combination with a novel therapeutic is to sustain reductions in severe malaria throughout childhood, our results suggest that these therapeutics will require either extended pre-liver stage duration or multi-stage activity.

Importantly, our findings highlight the public health value of a therapeutic with combined pre-liver and blood stage activity for malaria prevention in children. Model results showed that combining a multi-stage therapeutic with SMC led to greater reductions in cumulative severe cases throughout childhood than single-stage therapeutics. This population-level effect was not the result of any assumed individual-level synergy between intervention components but rather emerged from OpenMalaria’s mechanistic structure: each component acts independently on a different stage of the parasite life cycle. Specifically, blood stage activity may treat infections that escape pre-liver stage protection, preventing parasite densities from reaching thresholds associated with severe disease. This aligns with recent findings from the phase 2 trial of the blood stage vaccine candidate RH5.1, which reported greater efficacy against high-parasite density infections [36]. Deploying a preventive intervention with blood stage activity may also modify the development of blood stage immunity. Currently, the only approved malaria therapeutics with both liver and blood stage activity are oral antimalarial drugs such as SP-AQ. In the near future, this gap could be filled with the development of long-acting injectable formulation of an antimalarial drug that is active against multiple stages of the parasite life cycle [14–17]. In the longer term, we could expect substantial benefit from layering SMC with a combination vaccine such as RTS,S or R21 combined with an RH5-based vaccine candidate (ClinicalTrials.gov ID NCT05357560) [19], or monoclonal antibody.

Our results also suggest that combining a novel malaria therapeutic with SMC is likely to lead to the greatest public health benefits in settings where there are gaps in SMC’s effectiveness. A new vaccine, monoclonal antibody, or long-acting injectable can provide an additional layer of protection for children with poor adherence to SP-AQ, or who are absent for one or more SMC cycles. And, while we did not explicitly model resistance to SP-AQ, our results indicate that a novel therapeutic is likely to have its strongest benefits in regions where SP’s efficacy has been compromised by parasite resistance. A new therapeutic can also supplement SMC’s protection for the period between cycles when SP-AQ’s protection has waned or when the number of SMC cycles does not fully cover the malaria transmission season [37].

On the other hand, our results indicate that SMC may not need the support of a new therapeutic when coverage is already high. A longer pre-liver stage protection half-life was needed for a novel therapeutic to contribute additional reductions in malaria burden when children received five cycles of SMC. This result is consistent with previous modelling studies; modelling has indicated that synergisms between a pre-liver stage vaccine and mass drug administration are lost when coverage was high [38], and we have previously demonstrated that increasing SMC’s coverage through, for example, the deployment of a new oral chemoprevention drug can lead to substantial increases in SMC’s effectiveness [31].

These trade-offs between SMC’s coverage and the benefits of a partner therapeutic imply that the cost-effectiveness of a new therapeutic will depend on the public health effectiveness of existing interventions. The number of children missed by SMC varies across programs. For example, the ACCESS-SMC project found that SMC coverage varied between countries; as few as 24% of children in Chad and as many as 86% of children in Burkina Faso received all four cycles of SP-AQ [5]. We did not model cost-effectiveness outcomes in this work, due to the uncertainties inherent in costs for therapeutics that are still being developed. However, our results indicate that adding seasonal deployment of a pre-liver stage vaccine to SMC may not be cost-effective when SMC’s coverage has been optimised and the efficacy of SP-AQ remains high. Our results also highlight the important conceptual difference between evaluating an intervention with respect to its absolute number of malaria cases averted and relative to the standard of care; any assessment of intervention cost-effectiveness should be coupled with a strong understanding of the absolute malaria burden in a setting.

All modelling studies are limited by their assumptions. In this study, we measured impact on the cumulative incidence of uncomplicated and severe cases throughout childhood. These outcomes do not capture the potential benefits of malaria chemoprevention for non-malarial health outcomes, and depend strongly on our transmission model’s assumptions about the relationships between infection density, malaria immunity acquisition, and the development of clinical disease [23,24]. Future studies of intervention combinations with data from clinical trials and with alternative health outcomes will be important to confirm the findings of our work, and validate or support changes to model assumptions on anti-parasite and anti-disease immunity.

In addition, we made assumptions within our model structure to represent the dynamics of our malaria preventive therapeutics. The model structures used have been previously validated to data for pre-liver stage vaccines and blood stage antimalarial drugs, having been used to estimate the expected public health impact or RTS,S [29] and SMC [26,31]. However, to date, no blood stage vaccines, monoclonal antibodies, or long-acting injectable drugs have been approved by the World Health Organization for use in malaria prevention. Hence, there is limited data available to validate a model structure for the effects of a blood stage therapeutic; new modelling evidence will be needed as blood stage and multi-stage therapeutics advance through clinical trials. We also did not model transmission-blocking therapeutics, as SMC only targets young children and our outcomes focused on malaria burden in children under ten. As a result, the broader community-level benefits of a therapeutic with transmission-blocking activity would not be fully captured. Future modelling studies should assess the potential impact of multi-stage therapeutics with transmission-blocking activity, particularly in interventions targeting all age groups.

Finally, our estimates of benefit were based on outcomes simulated from a limited number of deployment modalities and endemic settings. In particular, we focused on evaluating the impact of combining a novel therapeutic with SMC in high transmission settings, where new tools are most needed to reduce the burden of malaria. Future modelling studies could explore combinations in perennial settings and for other use-cases and target age-groups. We also did not investigate how sex, gender, or ethnicity would influence our study outcomes, due to the limited data available for validating estimates of intervention effectiveness across these different groups. As a result of these limitations, our work does not offer forecasts of likely therapeutic impact when combined with a chemoprevention program. We instead aim to provide evidence to support investment decisions and inform candidate selection.

In conclusion, this study has provided quantitative evidence to articulate the benefits of new malaria prevention therapeutics during childhood. Our evidence provides funders and policymakers with new understandings of how interventions can be combined to provide added benefits over the standard of care before clinical trials. Results demonstrate the importance of prioritising co-deployment of a novel therapeutic in settings with limited coverage or effectiveness of SMC and deprioritising when the effectiveness of SMC has already been optimised. For the first time, our results demonstrate the potential for a multi-stage malaria therapeutic to improve a program’s ability to maintain benefits for children in the years after the deployment of an intervention has stopped.

## Author contributions

**Conceptualization:** Melissa A Penny, Lydia Braunack-Mayer

**Data Curation:** Lydia Braunack-Mayer

**Formal Analysis:** Lydia Braunack-Mayer

**Funding Acquisition:** Melissa A Penny

**Investigation:** Lydia Braunack-Mayer

**Methodology:** Melissa A Penny, Lydia Braunack-Mayer

**Project Administration:** Melissa A Penny, Sherrie L Kelly

**Resources:** Melissa A Penny

**Software:** Melissa A Penny, Lydia Braunack-Mayer, Josephine Malinga, Narimane Nekkab

**Supervision:** Melissa A Penny, Jörg J Möhrle

**Validation:** Melissa A Penny, Lydia Braunack-Mayer, Josephine Malinga, Narimane Nekkab

**Visualization:** Lydia Braunack-Mayer

**Writing – Original Draft Preparation:** Lydia Braunack-Mayer

**Writing – Review & Editing:** Lydia Braunack-Mayer, Josephine Malinga, Narimane Nekkab, Sherrie L Kelly, Jörg J Möhrle, Melissa A Penny

## Data Availability

All code and datasets are available on a GitHub repository at https://github.com/lydiab-mayer/modelling-malaria-combination-interventions/releases/tag/v1.0.0.

## Acknowledgements

We wish to thank Dr Jean-Luc Bodmer (Gates Foundation) for his contribution to this project. We would also like to thank Dr Thiery Masserey for his project management support and acknowledge support and advice from all members of the Disease Modelling research unit of the Swiss Tropical and Public Health Institute. Calculations were performed at sciCORE (http://scicore.unibas.ch/) scientific computing centre at the University of Basel.

## Supplement to

## 1. Methods

### Malaria transmission model

We used an established individual-based model of malaria transmission, OpenMalaria, to evaluate the benefits of combining seasonal deployment of a novel medical prevention intervention with seasonal malaria chemoprevention (SMC). OpenMalaria was originally developed to estimate the potential public-health impact of a malaria vaccine [1]. This stochastic model is open source (https://github.com/SwissTPH/openmalaria/wiki) and has been described previously [1,2]. Our study used OpenMalaria version 45 (https://swisstph.github.io/openmalaria/schema-45.html).

In table 1.1, we provide a summary of the key model dynamics relevant for this study.

**Table 1.1:**
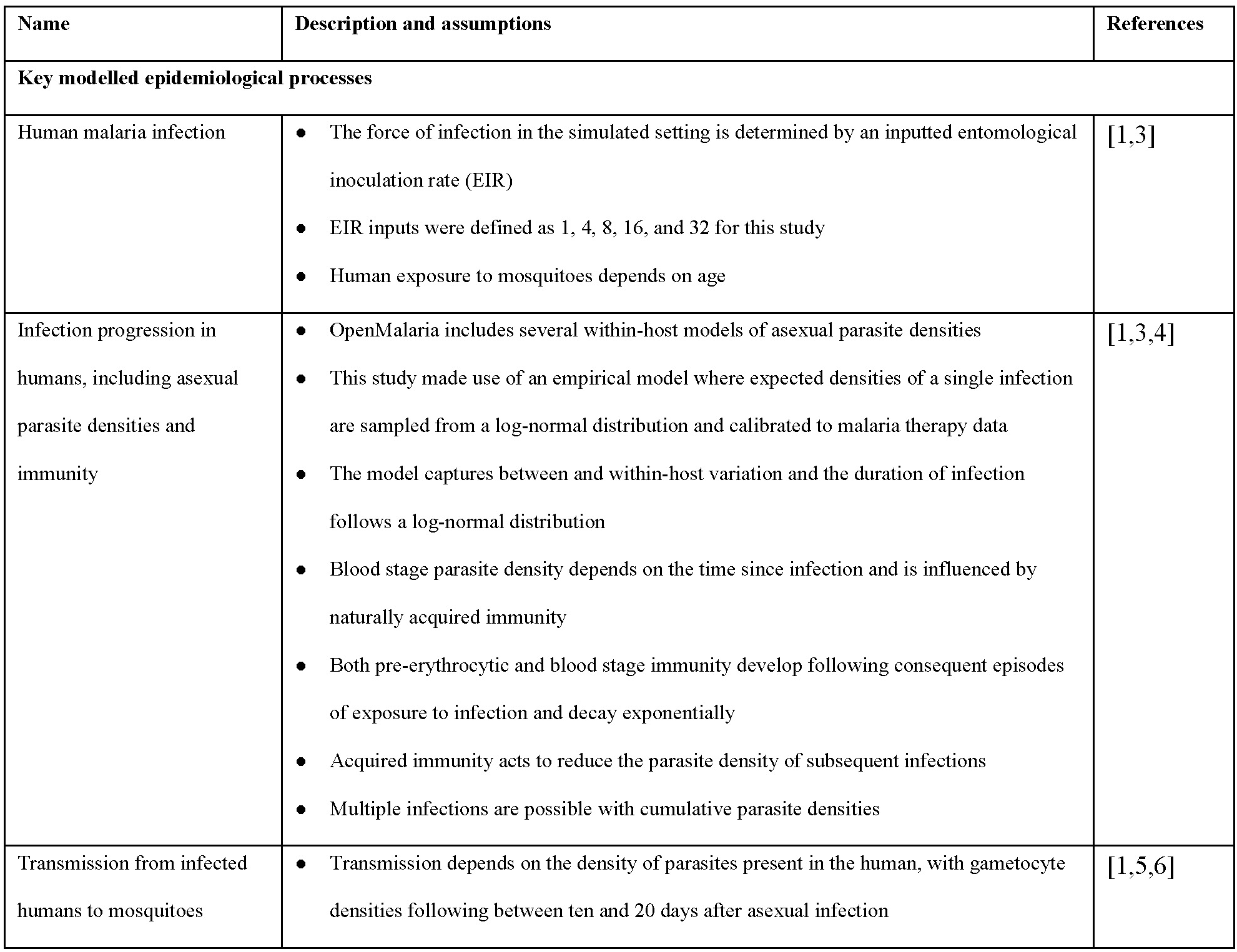
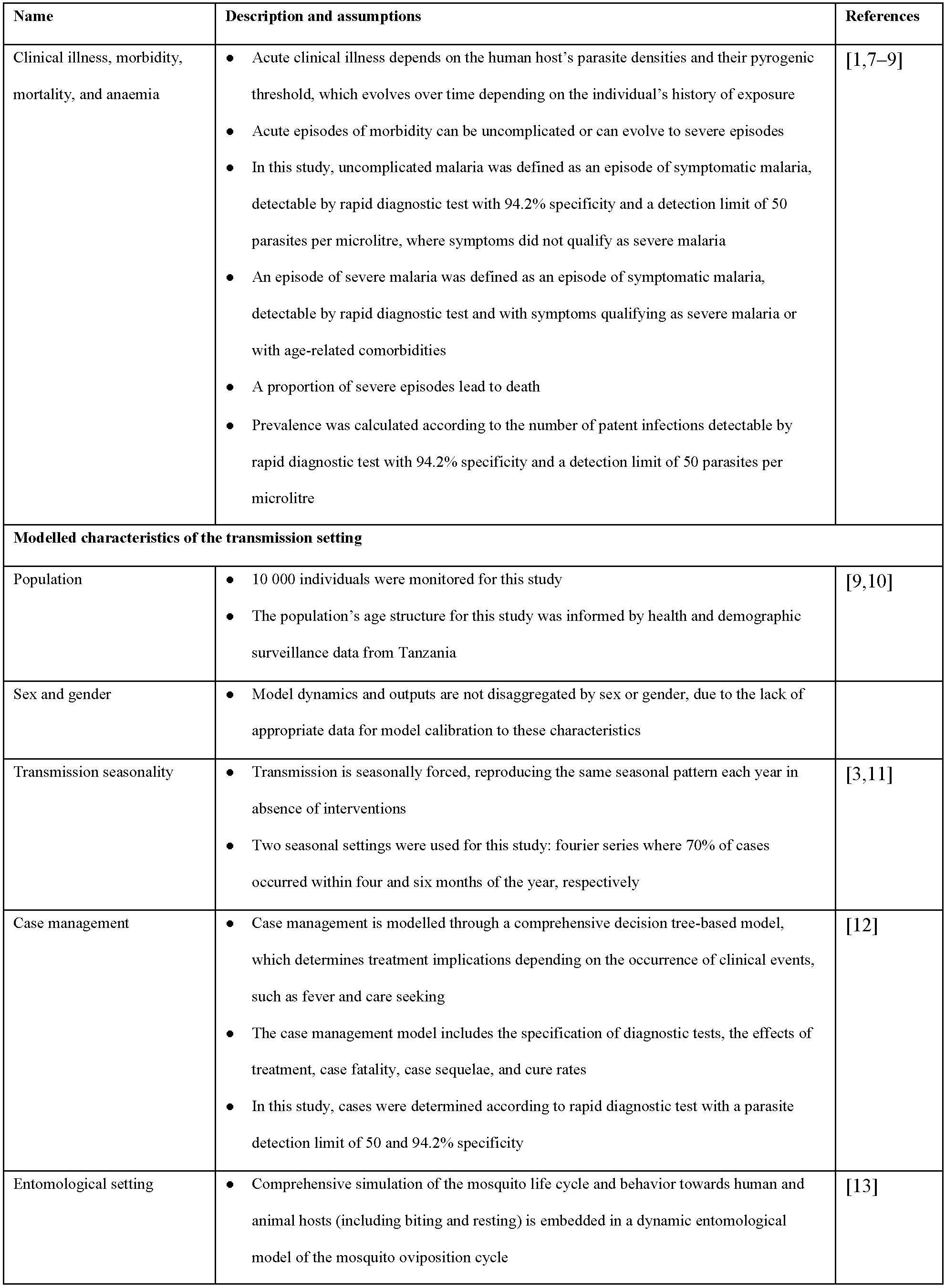
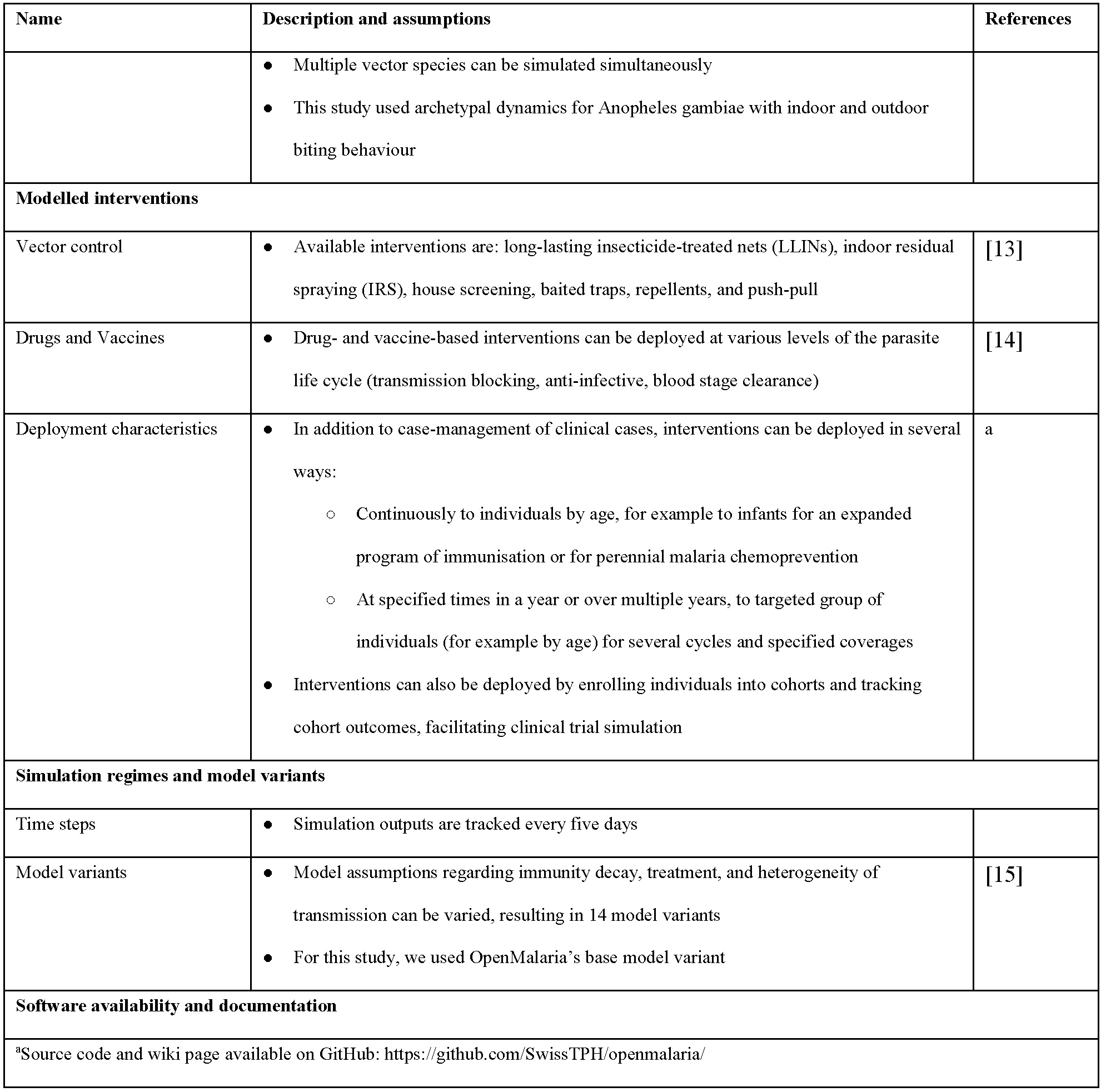
Summary of disease model characteristics, adapted from earlier OpenMalaria publications [2,16,17].

| Name | Description and assumptions | References |
| --- | --- | --- |
| **Key modelled epidemiological processes** |  |  |
| Human malaria infection | The force of infection in the simulated setting is determined by an inputted entomological inoculation rate (EIR)<br>EIR inputs were defined as 1, 4, 8, 16, and 32 for this study<br>Human exposure to mosquitoes depends on age | [1,3] |
| Infection progression in humans, including asexual parasite densities and immunity | OpenMalaria includes several within-host models of asexual parasite densities<br>This study made use of an empirical model where expected densities of a single infection are sampled from a log-normal distribution and calibrated to malaria therapy data<br>The model captures between and within-host variation and the duration of infection follows a log-normal distribution<br>Blood stage parasite density depends on the time since infection and is influenced by naturally acquired immunity<br>Both pre-erythrocytic and blood stage immunity develop following consequent episodes of exposure to infection and decay exponentially<br>Acquired immunity acts to reduce the parasite density of subsequent infections<br>Multiple infections are possible with cumulative parasite densities | [1,3,4] |
| Transmission from infected humans to mosquitoes | Transmission depends on the density of parasites present in the human, with gametocyte densities following between ten and 20 days after asexual infection | [1,5,6] |
| Clinical illness, morbidity, mortality, and anaemia | Acute clinical illness depends on the human host's parasite densities and their pyrogenic threshold, which evolves over time depending on the individual's history of exposure<br>Acute episodes of morbidity can be uncomplicated or can evolve to severe episodes<br>In this study, uncomplicated malaria was defined as an episode of symptomatic malaria, detectable by rapid diagnostic test with 94.2% specificity and a detection limit of 50 parasites per microlitre, where symptoms did not qualify as severe malaria<br>An episode of severe malaria was defined as an episode of symptomatic malaria, detectable by rapid diagnostic test and with symptoms qualifying as severe malaria or with age-related comorbidities<br>A proportion of severe episodes lead to death<br>Prevalence was calculated according to the number of patent infections detectable by rapid diagnostic test with 94.2% specificity and a detection limit of 50 parasites per microlitre | [1,7–9] |
| **Modelled characteristics of the transmission setting** |  |  |
| Population | 10 000 individuals were monitored for this study<br>The population's age structure for this study was informed by health and demographic surveillance data from Tanzania | [9,10] |
| Sex and gender | Model dynamics and outputs are not disaggregated by sex or gender, due to the lack of appropriate data for model calibration to these characteristics |  |
| Transmission seasonality | Transmission is seasonally forced, reproducing the same seasonal pattern each year in absence of interventions<br>Two seasonal settings were used for this study: fourier series where 70% of cases occurred within four and six months of the year, respectively | [3,11] |
| Case management | Case management is modelled through a comprehensive decision tree-based model, which determines treatment implications depending on the occurrence of clinical events, such as fever and care seeking<br>The case management model includes the specification of diagnostic tests, the effects of treatment, case fatality, case sequelae, and cure rates<br>In this study, cases were determined according to rapid diagnostic test with a parasite detection limit of 50 and 94.2% specificity | [12] |
| Entomological setting | Comprehensive simulation of the mosquito life cycle and behavior towards human and animal hosts (including biting and resting) is embedded in a dynamic entomological model of the mosquito oviposition cycle | [13] |
|  | Multiple vector species can be simulated simultaneously<br>This study used archetypal dynamics for *Anopheles gambiae* with indoor and outdoor biting behaviour |  |
| **Modelled interventions** |  |  |
| Vector control | Available interventions are: long-lasting insecticide-treated nets (LLINs), indoor residual spraying (IRS), house screening, baited traps, repellents, and push-pull | [13] |
| Drugs and Vaccines | Drug- and vaccine-based interventions can be deployed at various levels of the parasite life cycle (transmission blocking, anti-infective, blood stage clearance) | [14] |
| Deployment characteristics | In addition to case-management of clinical cases, interventions can be deployed in several ways:<br>Continuously to individuals by age, for example to infants for an expanded program of immunisation or for perennial malaria chemoprevention<br>At specified times in a year or over multiple years, to targeted group of individuals (for example by age) for several cycles and specified coverages<br>Interventions can also be deployed by enrolling individuals into cohorts and tracking cohort outcomes, facilitating clinical trial simulation | a |
| **Simulation regimes and model variants** |  |  |
| Time steps | Simulation outputs are tracked every five days |  |
| Model variants | Model assumptions regarding immunity decay, treatment, and heterogeneity of transmission can be varied, resulting in 14 model variants<br>For this study, we used OpenMalaria's base model variant | [15] |
| **Software availability and documentation** |  |  |
| *Source code and wiki page available on GitHub: https://github.com/SwissTPH/openmalaria/ |  |  |

### Outcome measures

To evaluate the benefits of adding a new therapeutic to an existing malaria prevention program, we measured reductions in the cumulative incidence of uncomplicated and severe malaria throughout childhood. The calculations for these metrics involved the following steps:

1. Using OpenMalaria, we simulated disease outcomes for a cohort of children born in the first year of intervention delivery for a ten-year period.
2. We evaluated the cumulative incidence of uncomplicated and severe malaria cases when children reached five and ten years old.
3. As illustrated in figure 1, we captured these cumulative incidences under three scenarios: children received only SMC, children received only the novel therapeutic, and children received both SMC and the novel therapeutic.
4. The protective efficacy of the combined intervention was then calculated relative to two counterfactual scenarios, as

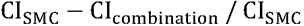

or as

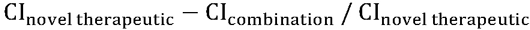

for CI_SMC_ the cumulative incidence of uncomplicated or severe malaria cases by five or ten years old for children receiving only SMC, CI_novel therapeutic_ the cumulative incidence for children receiving only the novel therapeutic, and CI_combination_ the cumulative incidence for children receiving the combined intervention.

These relative measures of impact were chosen because they capture the likely benefits of a seasonally deployed intervention as it would be measured in phase three or four clinical trials. For example, the R21/Matrix-M phase three clinical trial evaluated vaccine efficacy relative to a control arm where children received regular deployment of SMC [18]. The phase three trial of seasonal vaccination with RTS,S/AS01E with or without SMC also reported the protective efficacy of the combined interventions relative to both children who received only RTS,S/AS01E and only SMC [19].

### Intervention dynamics and deployment

Key model parameters included the component’s initial efficacy at the time of administration, the protection half-life or number of days until therapeutic efficacy reached half of its initial efficacy, and the shape of efficacy decay.

**Figure 1.1.**
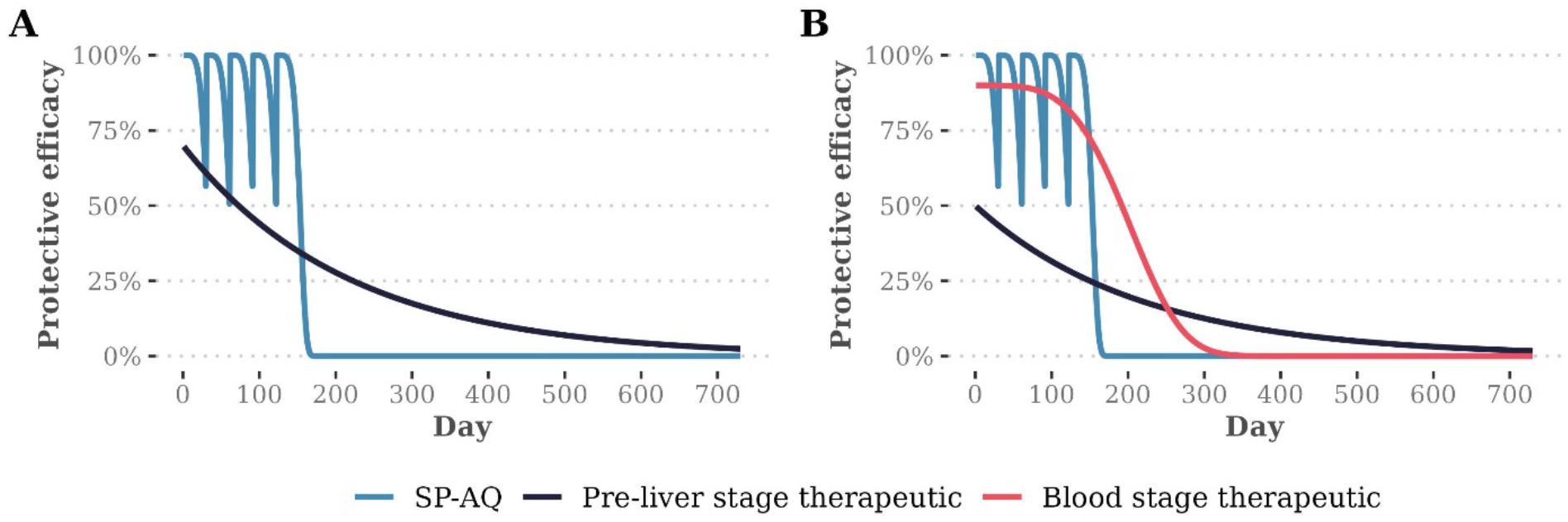
Illustration of key model parameters

**Figure 1.2:**
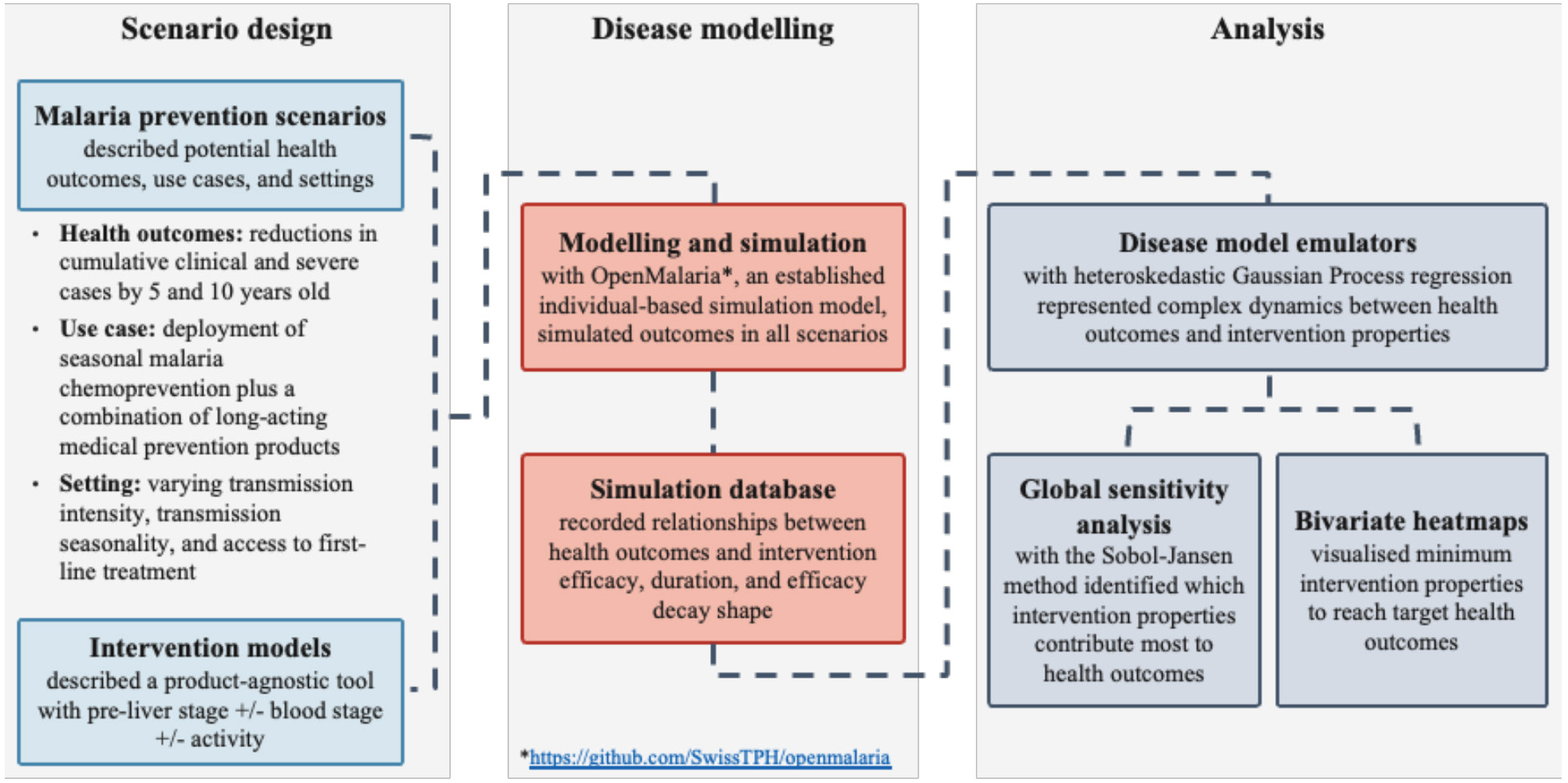
Schematic diagram of analysis framework for defining essential product characteristics for malaria prevention interventions

Initial efficacy refers to the therapeutic’s effect at time of administration as a percentage of the maximum possible effect. Protection half-life is the number of days until the effect decays to 50% of its initial efficacy. This parameter value is the scale parameter used in the Weibull function for effect decay. The decay shape is the shape parameter used in the Weibull function for effect decay. Panel A illustrates five cycles of SMC with SP-AQ together with a pre-liver stage therapeutic with 70% initial efficacy, a protection half-life of 150 days, and a long tail of protection (decay shape parameter of 1). Panel B illustrates five cycles of SMC with SP-AQ together with a multi-stage therapeutic with: 70% pre-liver stage initial efficacy, a pre-liver stage protection half-life of 150 days, a long pre-liver stage tail of protection (decay shape parameter of 1), 90% blood stage initial efficacy, a blood stage protection half-life of 200 days, and a rapid blood stage tail of protection (decay shape parameter of 4).

### Statistical analysis

Following our previously published methodology [16], and as illustrated in figure 1.2, we applied a predictive target product profile modelling framework to identify the therapeutic properties associated with maximum public health benefit when combining SMC with novel interventions.

We began by generating simulations from the OpenMalaria individual-based transmission model across a discrete sample of input intervention parameters. A total of 500 parameter sets were generated using Latin hypercube sampling with a uniform probability distribution, ensuring uniform coverage of a multidimensional input space. Parameter sets were sampled from the ranges shown in table 1, which were chosen to represent the broadest possible space of likely properties of a next-generation prevention therapeutic. For each sampled intervention profile, defined by the values for initial efficacy, protection half-life, and decay shape, we simulated ten stochastic replicates to capture stochastic variation in model outcomes.

Next, we trained a heteroskedastic Gaussian Process regression model using the hetGP package in R [20] to emulate the relationship between input intervention parameters and simulated health outcomes (i.e., cumulative incidence of uncomplicated and severe malaria). This surrogate model, known as a model emulator, approximates the complex and computationally intensive dynamics of the individual-based model without needing to perform a full simulation, allowing computationally efficient evaluation across a broader parameter space. Emulators were trained separately for each model scenario (type of therapeutic, seasonality profile, level of access to care and deployment scenario).

Gaussian Process regression models have previously been used successfully to emulate therapeutic dynamics in OpenMalaria in this exact framework [16,17,21]. Accordingly, we did not perform additional cross-validation for model selection. Instead, we assessed emulator performance using the R-squared correlation coefficient between observed and predicted values on a 10% hold-out validation set. To further confirm model fit, we examined both the R-squared values and scatter plots of true versus predicted outcomes.

To assess the influence of each intervention property on model outcomes, we conducted a nonparametric variance-based sensitivity analysis using the sensitivity package in R [22]. We applied the Sobol-Jansen method [23] to estimate Sobol total-order indices, which quantify the contribution of each input parameter, including all interaction effects, to the variance of the output. This analysis was performed on two large input datasets (50,000 parameter samples each) generated via Latin hypercube sampling, with 1000 bootstrap replicates used to estimate uncertainty.

Finally, to identify minimum required therapeutic properties for achieving target health outcomes, we used the trained emulators to create bivariate heatmaps. For each model scenario, emulators were used to predict health outcomes for a grid of therapeutic properties (e.g., protection half-life, initial efficacy). The predictions were used to generate heatmaps, allowing us to identify the minimum value of each property (e.g., protection half-life) for which the mean prediction remained above a chosen public health target (e.g., ≥10% reduction in cumulative severe malaria by age 10).

All statistical analyses were conducted in R (version 4.3.2) [24]. Calculations were performed at sciCORE (http://scicore.unibas.ch/) scientific computing centre at the University of Basel.

## 2. Additional results: Combining SMC with seasonal deployment of a pre-liver stage product

The median number of cumulative cases per year of age were calculated when children aged three to 59 months received one of the following interventions: no intervention, five SMC cycles (perfect deployment scenario), seasonal deployment of a pre-liver stage therapeutic, or the combination of five SMC cycles and the pre-liver stage therapeutic. Top panels include deployment of a pre-liver stage therapeutic with a protection half-life of 354 days, 90% initial efficacy, and decay shape parameter of 1. Bottom panels include a pre-liver stage therapeutic with a protection half-life of 120 days, 50% initial efficacy, and decay shape parameter of 1. The pink shaded regions indicate age-eligibility for SMC and the novel therapeutic. Shaded regions indicate the minimum and maximum cumulative cases per person observed across stochastic replicates of simulations from the individual-based malaria transmission model. Model results are shown for a high transmission scenario (32% *Pf*PR_2-10_) where 75% of malaria cases occur within six months of the year and the probability of seeking first-line treatment for clinical malaria over 14 days is low (10%).

**Figure 2.1:**
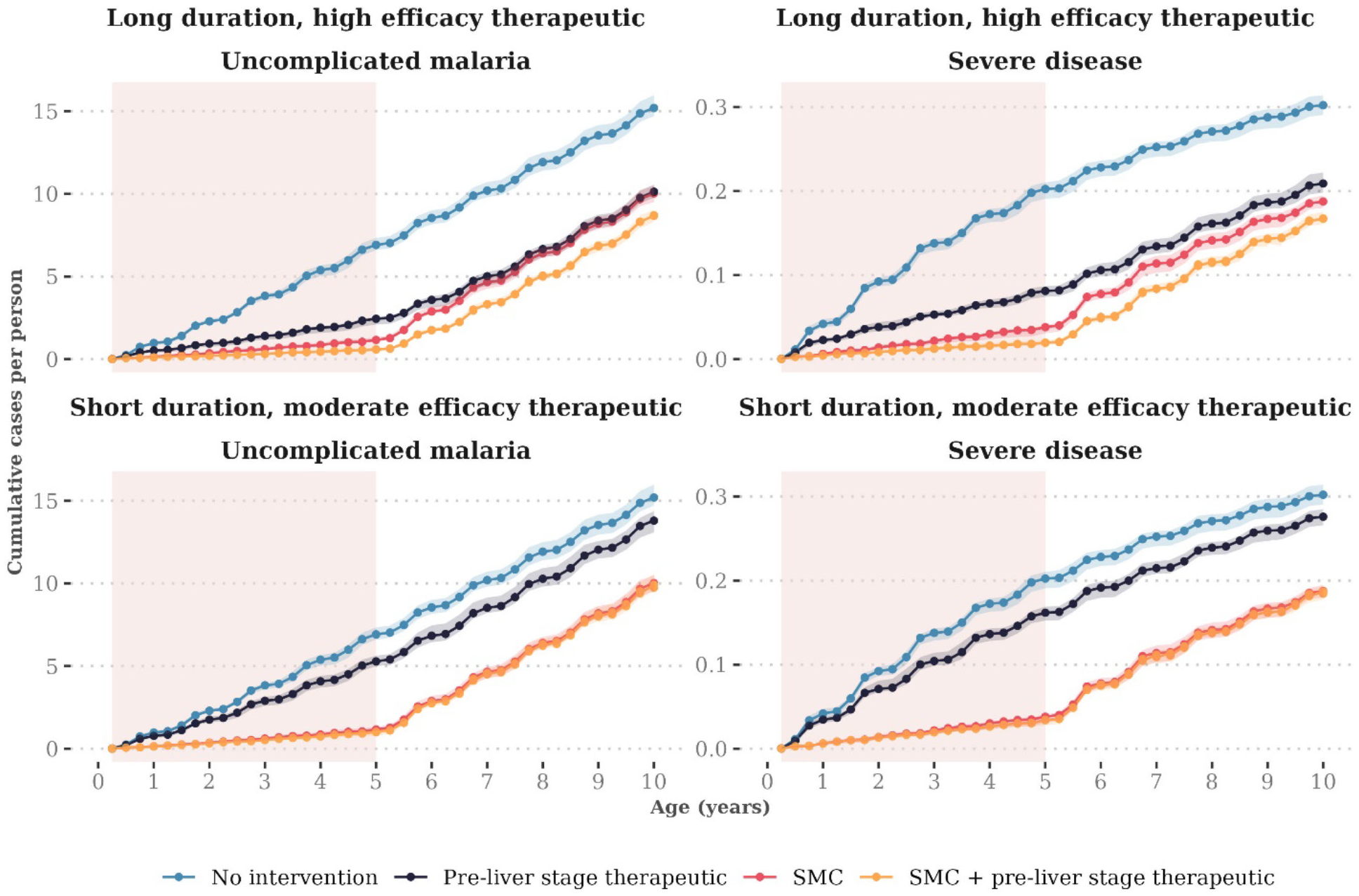
Predicted cumulative case curves when seasonal deployment of two exemplar pre-liver stage products are combined with perfect deployment of SMC

The median number of cumulative cases per year of age were calculated when children aged three to 59 months received one of the following interventions: no intervention, four SMC cycles (imperfect seasonal coverage scenario), seasonal deployment of a pre-liver stage therapeutic, or the combination of four SMC cycles and the pre-liver stage therapeutic. Top panels include deployment of a pre-liver stage therapeutic with a protection half-life of 354 days, 90% initial efficacy, and decay shape parameter of 1. Bottom panels include a pre-liver stage therapeutic with a protection half-life of 120 days, 50% initial efficacy, and decay shape parameter of 1. The pink shaded regions indicate age-eligibility for SMC and the novel therapeutic. Shaded regions indicate the minimum and maximum cumulative cases per person observed across stochastic replicates of simulations from the individual-based malaria transmission model. Model results are shown for a high transmission scenario (32% *Pf*PR_2-10_) where 75% of malaria cases occur within six months of the year and the probability of seeking first-line treatment for clinical malaria over 14 days is low (10%).

Results show the imperfect deployment scenario, where children received three SMC cycles. Transmission is high (32% *Pf*PR_2-10_), 75% of malaria cases occur within six months of the year, and the probability of seeking first-line treatment for clinical malaria over 14 days is low (10%). Each panel shows the median reduction in cumulative uncomplicated (solid lines) or severe cases (dashed lines) achieved by combining seasonal deployment of a pre-liver stage therapeutic with SMC, relative to cumulative cases when the pre-liver stage therapeutic is deployed alone.

**Figure 2.2:**
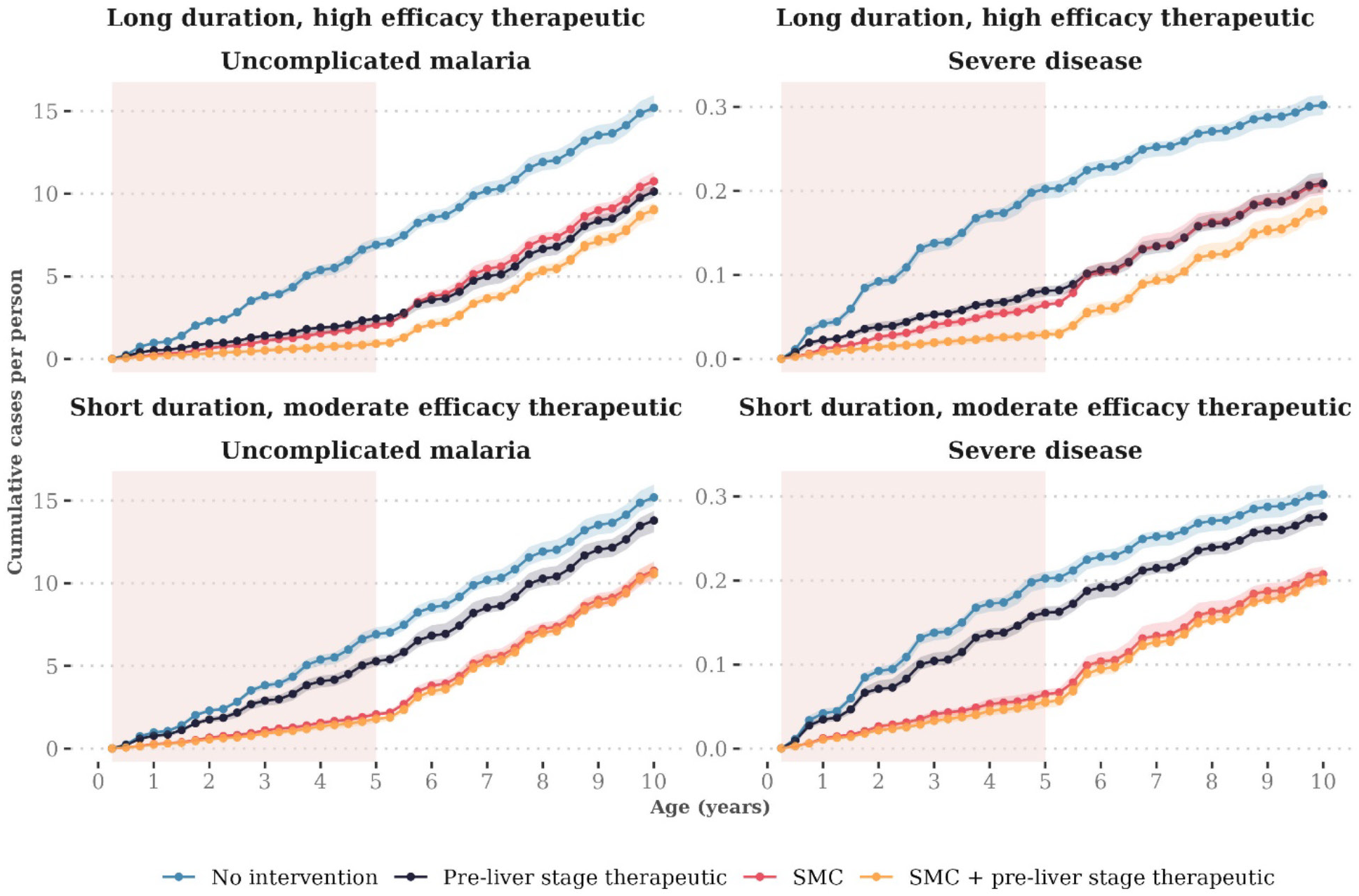
Predicted cumulative case curves when seasonal deployment of two exemplar pre-liver stage products are combined with imperfect seasonal coverage of SMC

Median reductions are calculated by dividing the parameter range for the therapeutic property shown on the x-axis (protection half-life, initial efficacy, decay shape) into 51 segments, and calculating the median outcome for each given segment across all other parameter values. Shaded regions represent the 25% and 75% quantiles of the corresponding reductions. Emulator predictions are shown separately for cumulative case outcomes at five (panels A) and ten years old (panels B). For outcomes at five, reductions of approximately 65% were observed across the full range of pre-liver stage parameter values, which is similar to the protective efficacy observed for the combination of RTS,S and SMC relative to SMC alone (59·0%, 95% CI 54·7 to 62·8).[19] This near-constant additional benefit is likely because SMC provides a curative antimalarial drug and is thus able to clear infections that escape the pre-liver stage product. The large size of the benefit is because we have evaluated SMC’s impact relative to the benefit of the pre-liver stage therapeutic deployed alone; in absolute terms, the additional number of cases averted by SMC will decrease with increasing efficacy and duration of the novel therapeutic.

**Figure 2.3:**
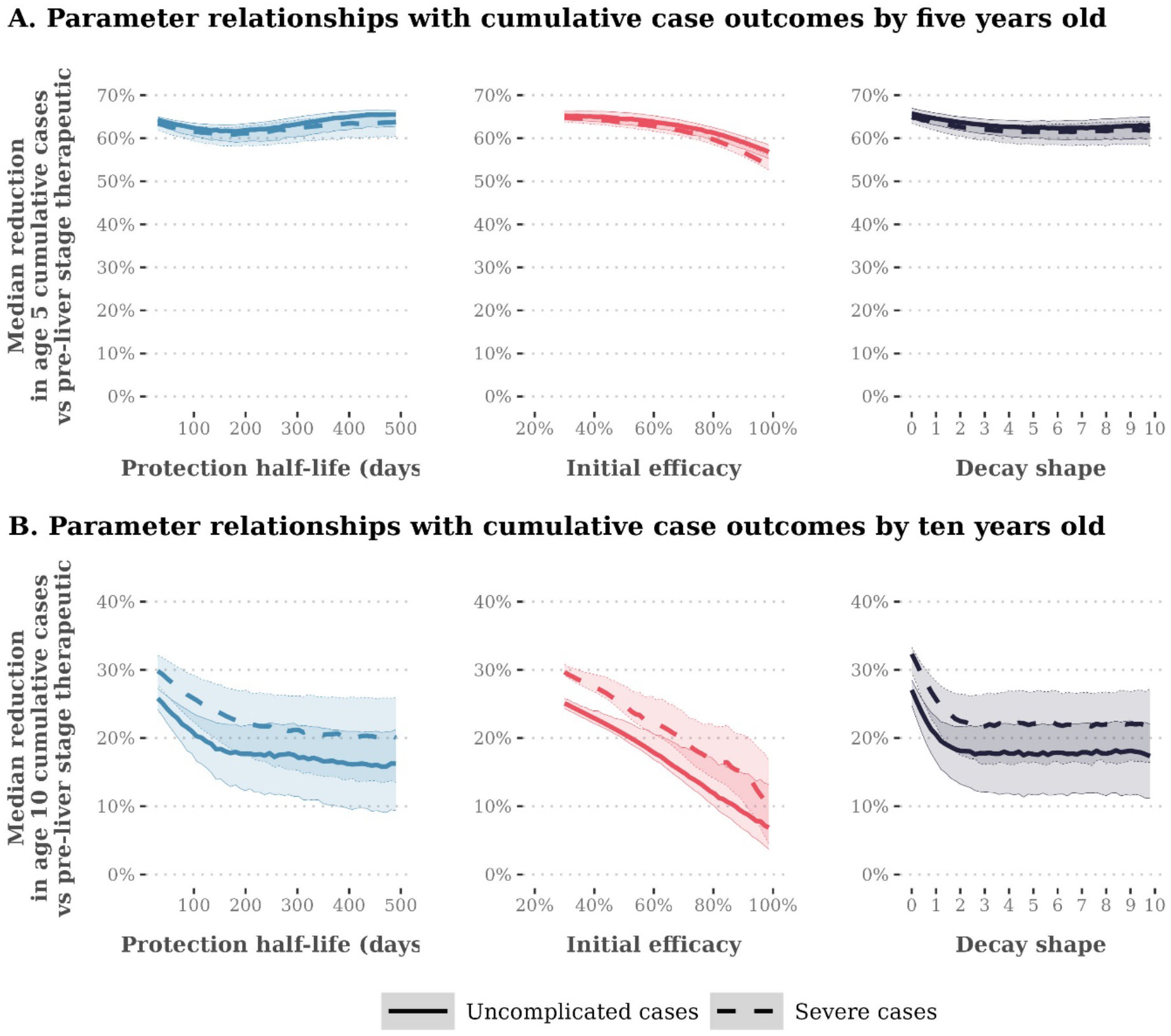
Gaussian Process regression emulator predictions for the relationship between pre-liver stage therapeutic properties and expected reductions in cumulative uncomplicated and severe cases when deployed with SMC, evaluated relative to seasonal deployment of a pre-liver stage therapeutic alone

Results show the perfect deployment scenario, where children aged three to 59 months received five SMC cycles. Transmission is high (32% *Pf*PR_2-10_), 75% of malaria cases occur within six months of the year, and the probability of seeking first-line treatment for clinical malaria over 14 days is low (10%). Each panel shows the median reduction in cumulative uncomplicated (solid lines) or severe cases (dashed lines) achieved by combining seasonal deployment of a pre-liver stage product with SMC, relative to cumulative cases when SMC is deployed alone.

Median reductions are calculated by dividing the parameter range for the product property shown on the x-axis (protection half-life, initial efficacy, decay shape) into 51 segments, and calculating the median outcome for each given segment across all other parameter values. Shaded regions represent the 25% and 75% quantiles of the corresponding reductions. Emulator predictions are shown separately for cumulative case outcomes at five (panels A) and ten years old (panels B).

**Figure 2.4:**
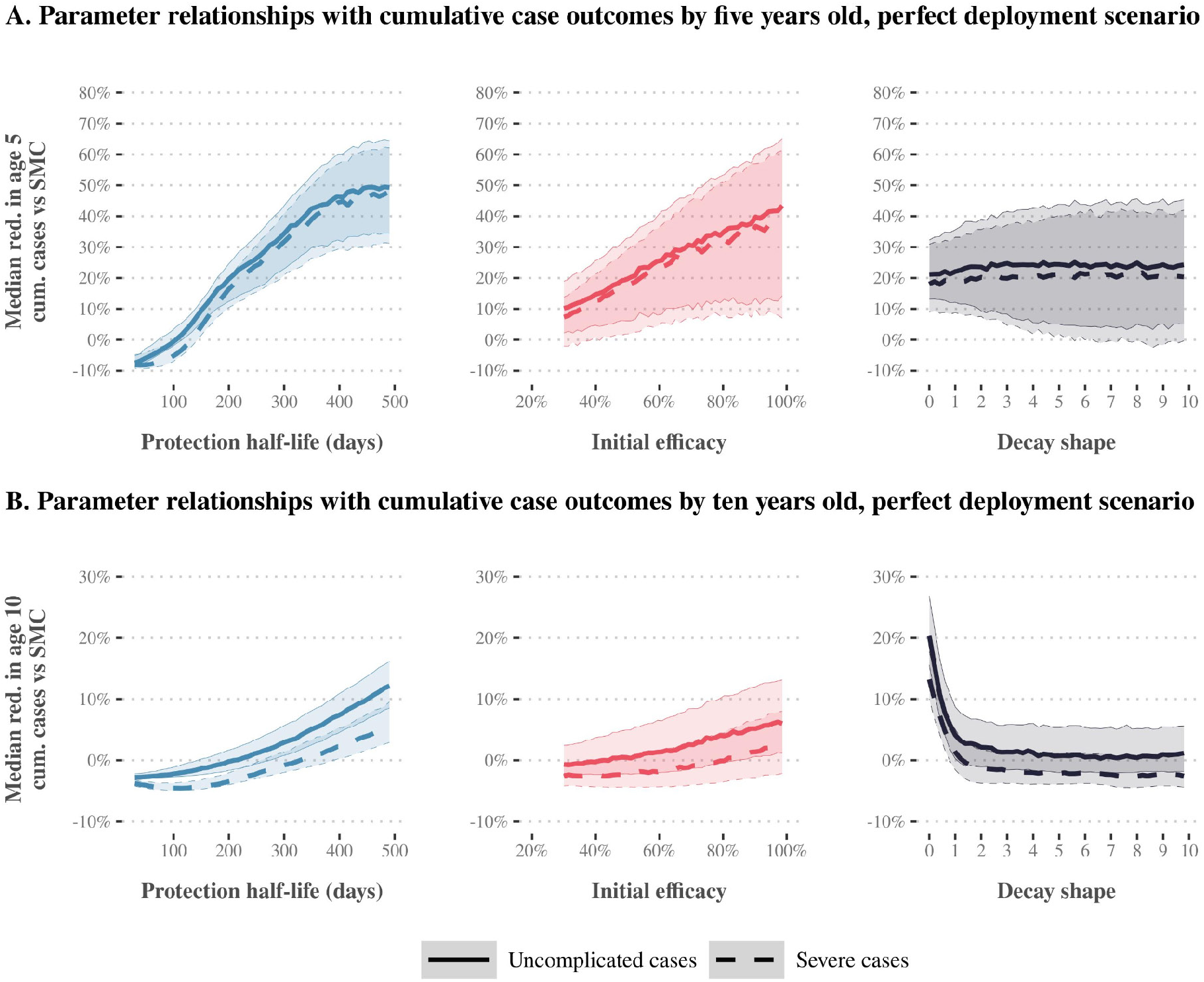
Gaussian Process regression emulator predictions for the relationship between pre-liver stage product properties and expected reductions in cumulative uncomplicated and severe cases relative to SMC alone, evaluated with perfect coverage of SMC

Results show the imperfect seasonal coverage scenario, where children aged three to 59 months received four SMC cycles. Transmission is high (32% *Pf*PR_2-10_), 75% of malaria cases occur within six months of the year, and the probability of seeking first-line treatment for clinical malaria over 14 days is low (10%). Each panel shows the median reduction in cumulative uncomplicated (solid lines) or severe cases (dashed lines) achieved by combining seasonal deployment of a pre-liver stage product with SMC, relative to cumulative cases when SMC is deployed alone. Median reductions are calculated by dividing the parameter range for the product property shown on the x-axis (protection half-life, initial efficacy, decay shape) into 51 segments, and calculating the median outcome for each given segment across all other parameter values. Shaded regions represent the 25% and 75% quantiles of the corresponding reductions. Emulator predictions are shown separately for cumulative case outcomes at five (panels A) and ten years old (panels B).

**Figure 2.5:**
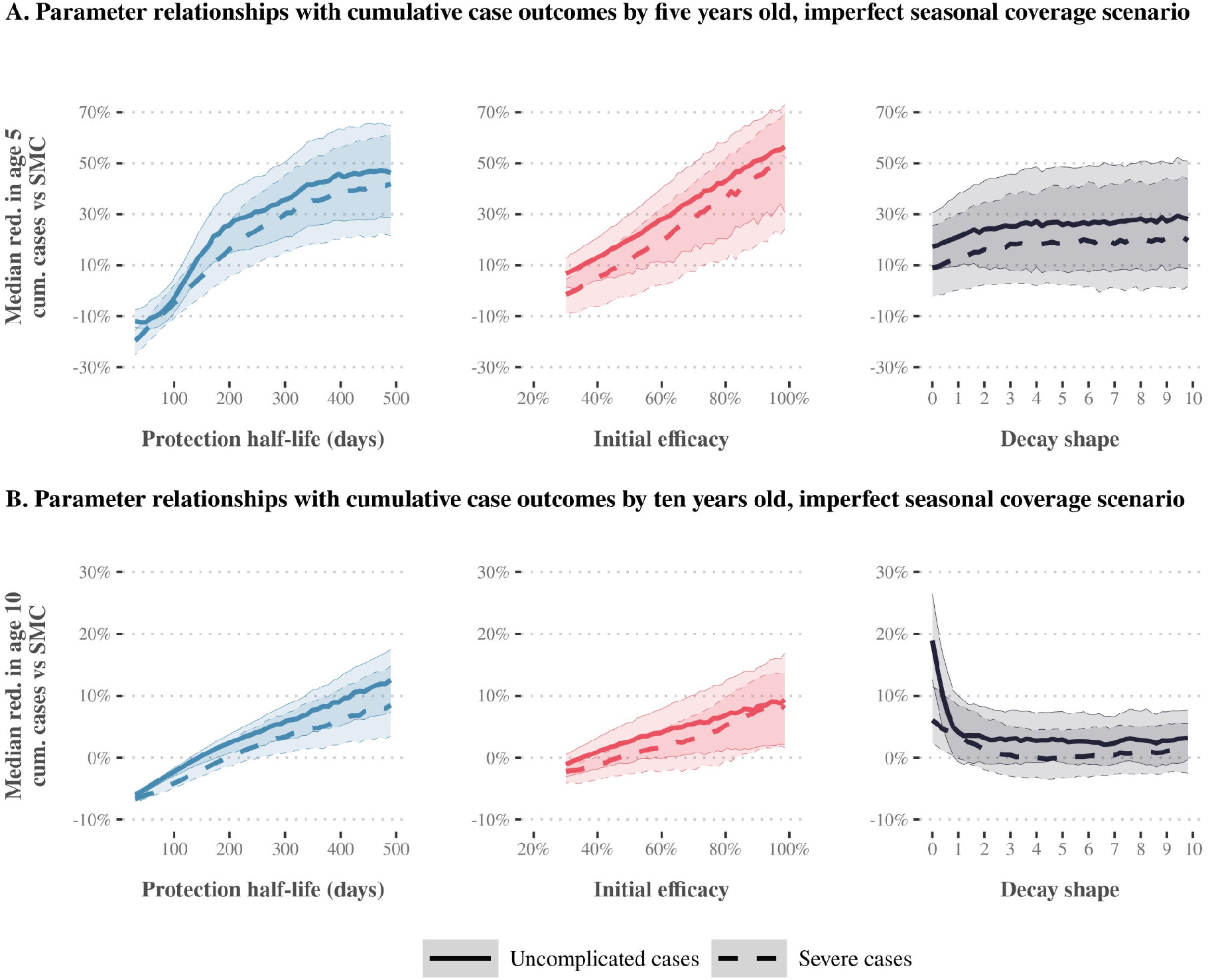
Gaussian Process regression emulator predictions for the relationship between pre-liver stage product properties and expected reductions in cumulative uncomplicated and severe cases relative to SMC alone, evaluated with imperfect seasonal coverage of SMC

Results show the random allocation scenario, where between 30% and 100% of children aged three to 59 months are randomly allocated to receive five SMC cycles. Transmission is high (32% *Pf*PR_2-10_), 75% of malaria cases occur within six months of the year, and the probability of seeking first-line treatment for clinical malaria over 14 days is low (10%). Each panel shows the median reduction in cumulative uncomplicated (solid lines) or severe cases (dashed lines) achieved by combining seasonal deployment of a pre-liver stage product with SMC, relative to cumulative cases when SMC is deployed alone. Median reductions are calculated by dividing the parameter range for the product property shown on the x-axis (protection half-life, initial efficacy, decay shape) into 51 segments, and calculating the median outcome for each given segment across all other parameter values. Shaded regions represent the 25% and 75% quantiles of the corresponding reductions. Emulator predictions are shown separately for cumulative case outcomes at five (panels A) and ten years old (panels B).

**Figure 2.6:**
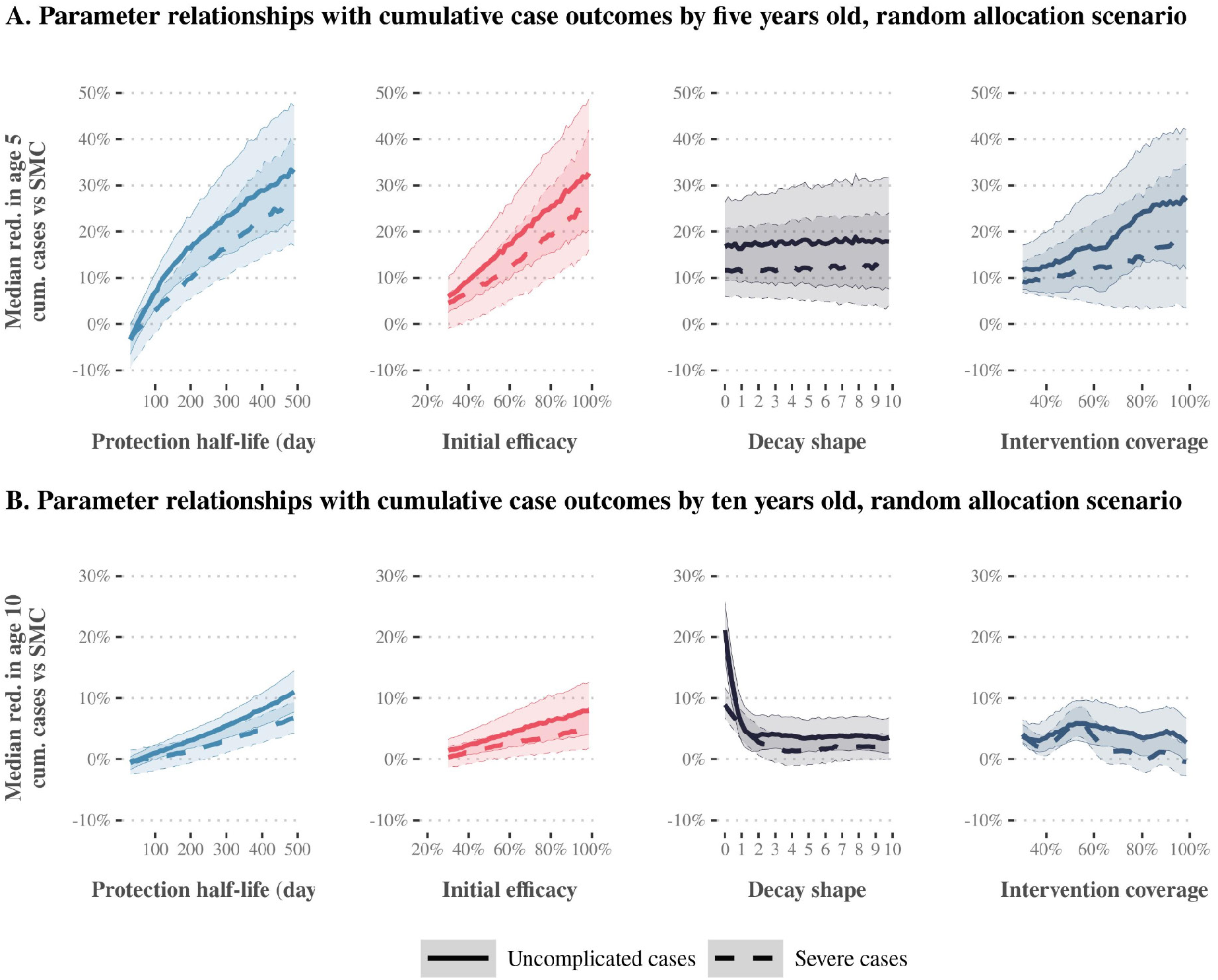
Gaussian Process regression emulator predictions for the relationship between pre-liver stage product properties and expected reductions in cumulative uncomplicated and severe cases relative to SMC alone, evaluated with random allocation of SMC cycles

Each square in each grid indicates the predicted reduction if a pre-liver stage product with the given protection half-life (x-axis) and initial efficacy (y-axis) were deployed in addition to SMC, assuming a long tail of protection (decay shape parameter 0.7). Reductions are rounded to the nearest 5%. Results show a scenario where transmission is high (ranging between 19% and 33% *Pf*PR_2-10_ depending on the seasonality profile and level of access to care). Panels depict predicted relationships for four different SMC deployment scenarios, two different seasonality profiles, and two different levels of access to first-line treatment for clinical malaria over 14 days (access to care).

**Figure 2.7:**
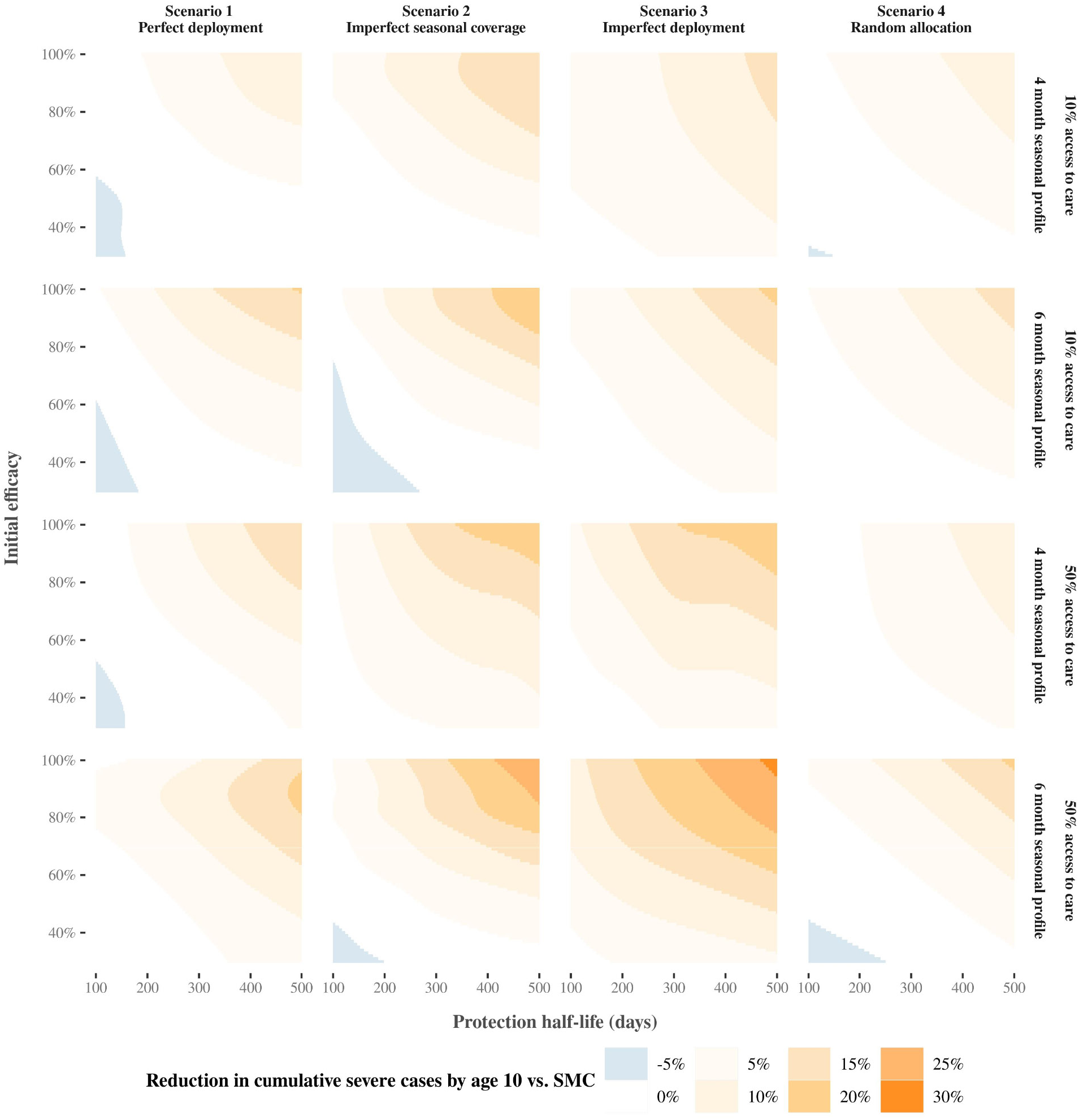
Predicted relationships between pre-liver stage protection half-life, initial efficacy and reductions in cumulative severe cases by ten years old, compared across settings and deployment scenarios for a product with a long tail of protection

Each square in each grid indicates the predicted reduction if a pre-liver stage product with the given protection half-life (x-axis) and initial efficacy (y-axis) were deployed in addition to SMC, assuming a rapid decay in protection (decay shape parameter 4). Reductions are rounded to the nearest 5%. Results show a scenario where transmission is high (32% *Pf*PR_2-10_), 75% of malaria cases occur within six months of the year, and the probability of seeking first-line treatment for clinical malaria over 14 days is low (10%). Panels depict predicted relationships for four different SMC deployment scenarios.

**Figure 2.8:**
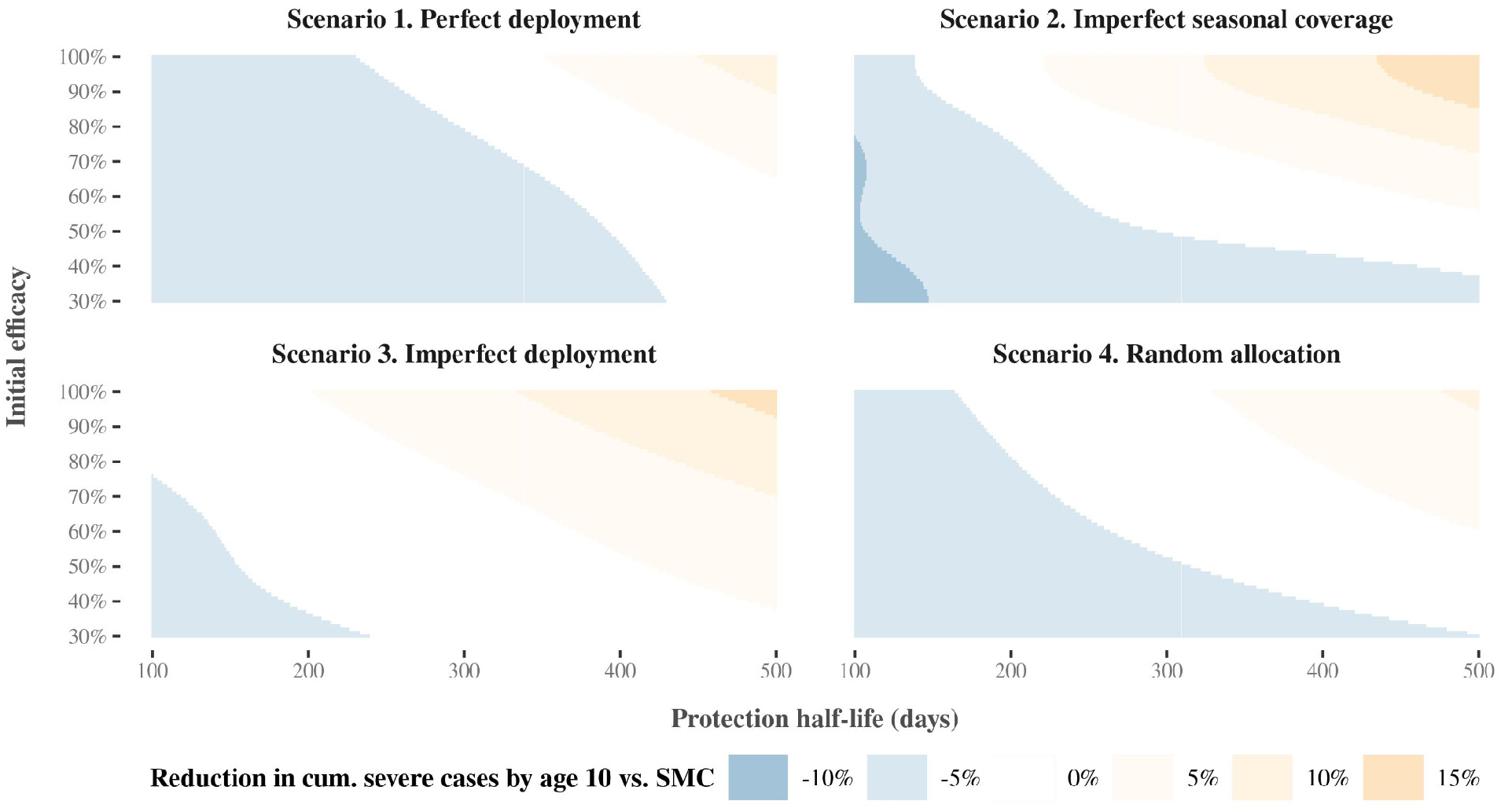
Predicted relationships between pre-liver stage protection half-life, initial efficacy and reductions in cumulative severe cases by ten years old, compared across SMC deployment scenarios for a product with a rapid decay in protection

## 3. Additional results: Combining SMC with seasonal deployment of a blood stage product

Results show the perfect deployment scenario, where children aged three to 59 months received five SMC cycles. Transmission is high (32% *Pf*PR_2-10_), 75% of malaria cases occur within six months of the year, and the probability of seeking first-line treatment for clinical malaria over 14 days is low (10%). Each panel shows the median reduction in cumulative uncomplicated (solid lines) or severe cases (dashed lines) achieved by combining seasonal deployment of a blood stage product with SMC, relative to cumulative cases when SMC is deployed alone. Median reductions are calculated by dividing the parameter range for the product property shown on the x-axis (protection half-life, initial efficacy, decay shape) into 51 segments, and calculating the median outcome for each given segment across all other parameter values. Shaded regions represent the 25% and 75% quantiles of the corresponding reductions. Emulator predictions are shown separately for cumulative case outcomes at five (panels A) and ten years old (panels B).

**Figure 3.1:**
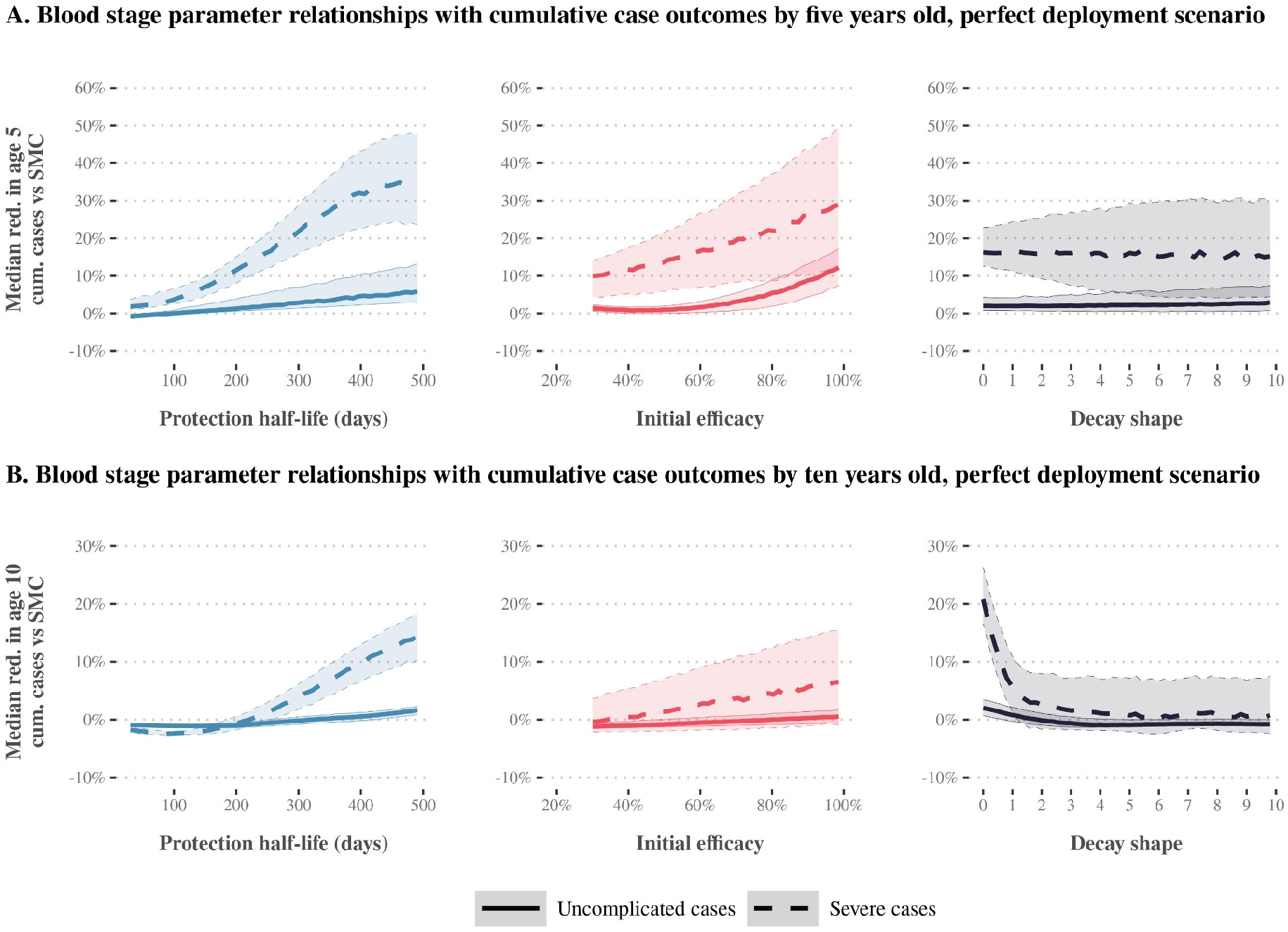
Gaussian Process regression emulator predictions for the relationship between blood stage product properties and expected reductions in cumulative uncomplicated and severe cases relative to SMC alone, evaluated with perfect coverage of SMC

Results show the imperfect seasonal coverage scenario, where children aged three to 59 months received four SMC cycles. Transmission is high (32% *Pf*PR_2-10_), 75% of malaria cases occur within six months of the year, and the probability of seeking first-line treatment for clinical malaria over 14 days is low (10%). Each panel shows the median reduction in cumulative uncomplicated (solid lines) or severe cases (dashed lines) achieved by combining seasonal deployment of a blood stage product with SMC, relative to cumulative cases when SMC is deployed alone. Median reductions are calculated by dividing the parameter range for the product property shown on the x-axis (protection half-life, initial efficacy, decay shape) into 51 segments, and calculating the median outcome for each given segment across all other parameter values. Shaded regions represent the 25% and 75% quantiles of the corresponding reductions. Emulator predictions are shown separately for cumulative case outcomes at five (panels A) and ten years old (panels B).

**Figure 3.2:**
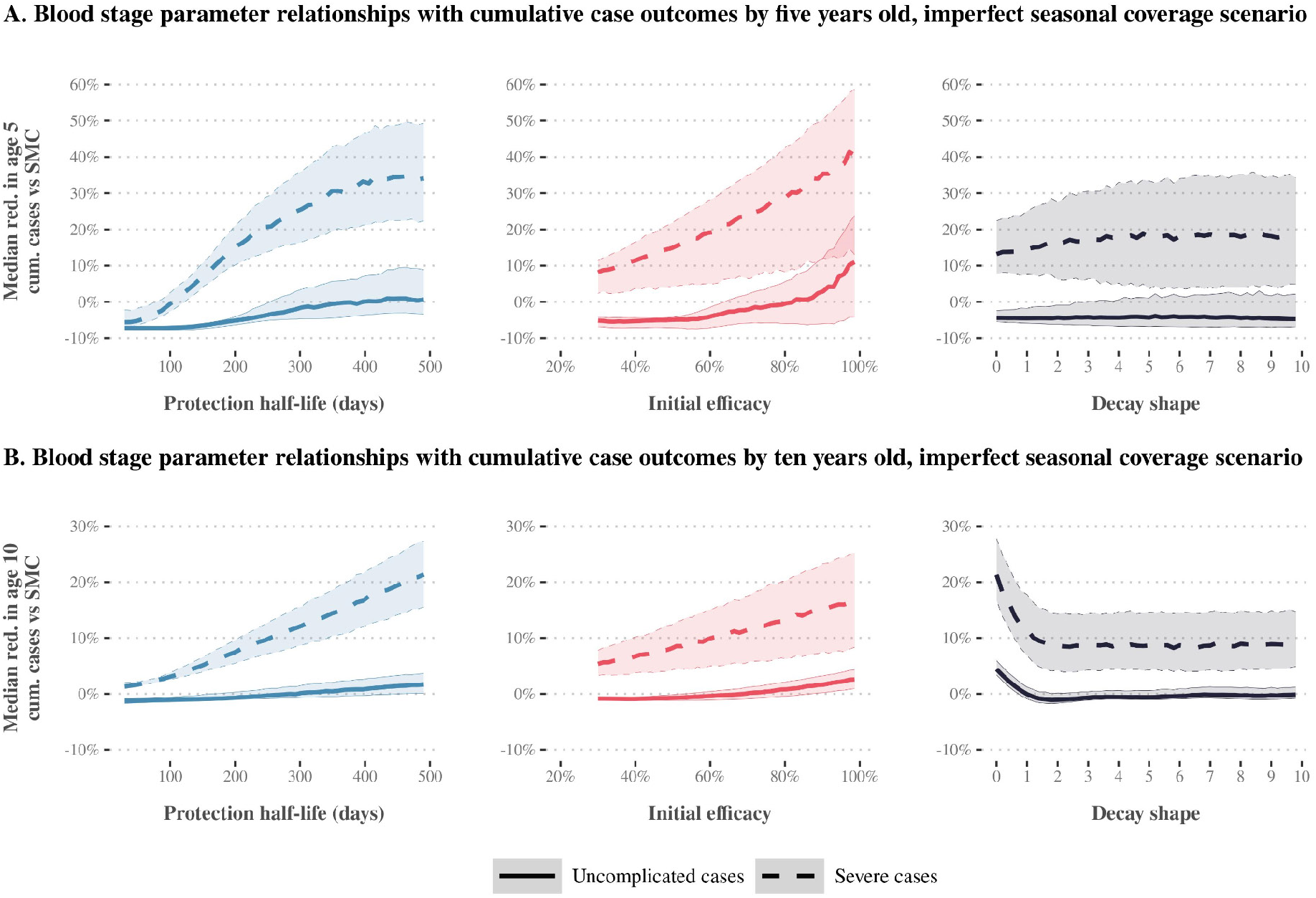
Gaussian Process regression emulator predictions for the relationship between blood stage product properties and expected reductions in cumulative uncomplicated and severe cases relative to SMC alone, evaluated with imperfect seasonal coverage of SMC

Results show the random allocation scenario, where between 30% and 100% of children aged three to 59 months are randomly allocated to receive five SMC cycles. Transmission is high (32% *Pf*PR_2-10_), 75% of malaria cases occur within six months of the year, and the probability of seeking first-line treatment for clinical malaria over 14 days is low (10%). Each panel shows the median reduction in cumulative uncomplicated (solid lines) or severe cases (dashed lines) achieved by combining seasonal deployment of a blood stage product with SMC, relative to cumulative cases when SMC is deployed alone. Median reductions are calculated by dividing the parameter range for the product property shown on the x-axis (protection half-life, initial efficacy, decay shape) into 51 segments, and calculating the median outcome for each given segment across all other parameter values. Shaded regions represent the 25% and 75% quantiles of the corresponding reductions. Emulator predictions are shown separately for cumulative case outcomes at five (panels A) and ten years old (panels B).

**Figure 3.3:**
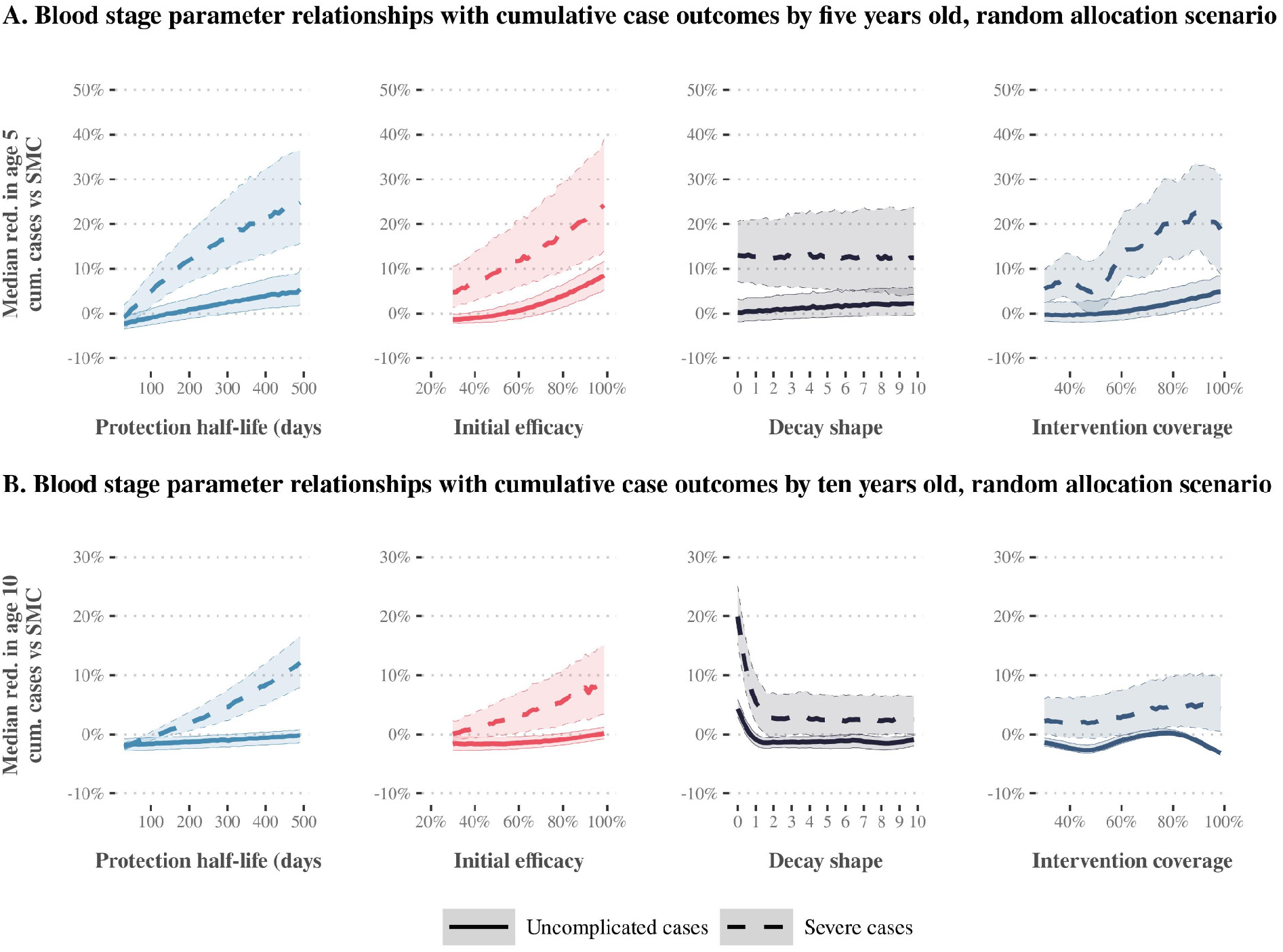
Gaussian Process regression emulator predictions for the relationship between blood stage product properties and expected reductions in cumulative uncomplicated and severe cases relative to SMC alone, evaluated with random allocation of SMC cycles

## 4. Additional results: Combining SMC with seasonal deployment of a multi-stage product

Bars show the total Sobol effect indices for intervention model parameters. Total Sobol effect indices can be interpreted as the proportion of variation in the cumulative case outcome that can be attributed to a small change in each model parameter and its interactions with other parameters. Indices are shown across levels of transmission intensity (x-axis) for a scenario where three yearly cycles of SMC are deployed (imperfect coverage scenario).

**Figure 4.1:**
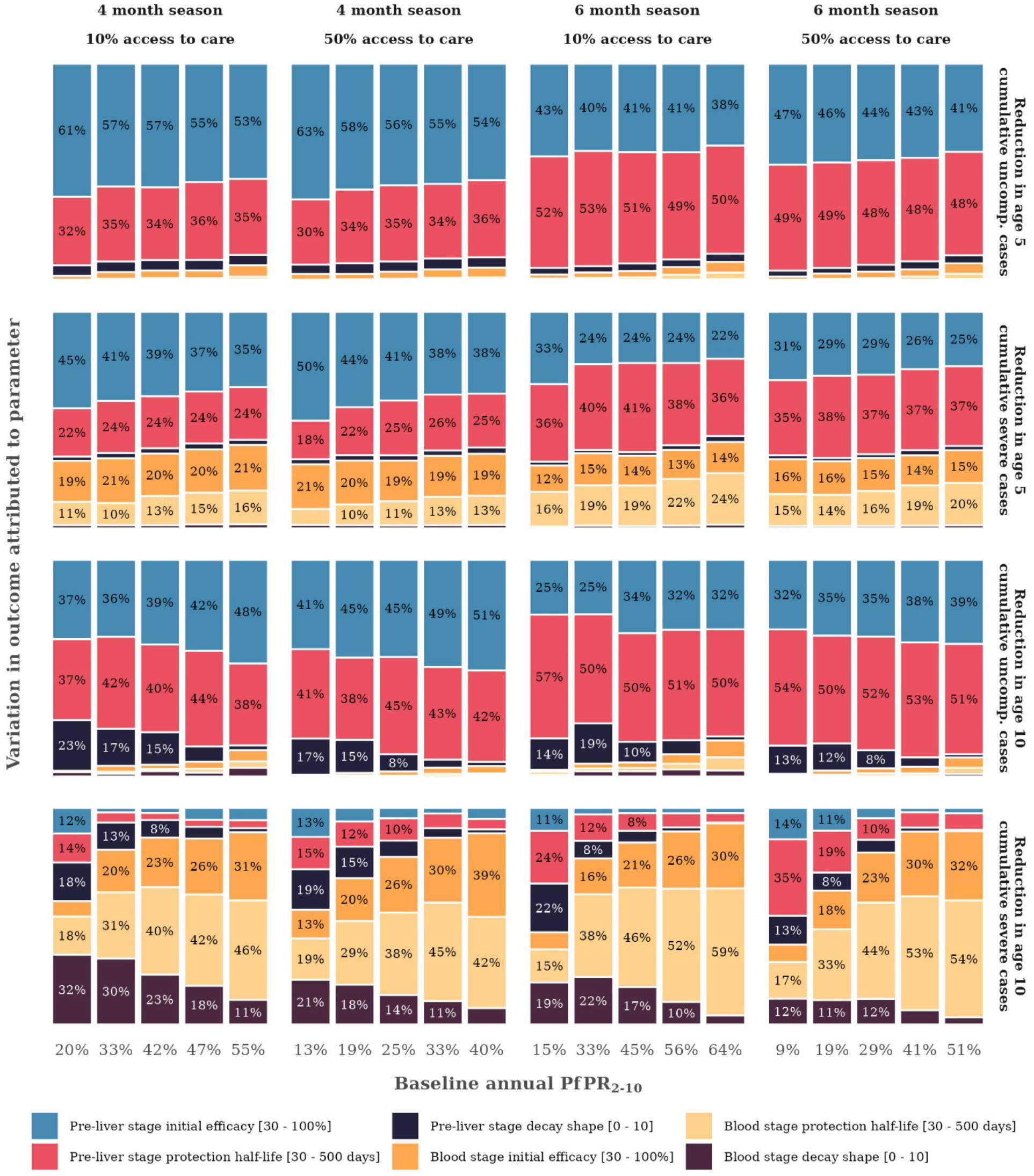
Drivers of impact on all cumulative case outcomes for a product with both pre-liver stage and blood stage activity deployed together with three yearly cycles of SMC, compared with SMC deployed alone

Results show the perfect deployment scenario, where children aged three to 59 months received five SMC cycles. 75% of malaria cases occur within six months of the year and the probability of seeking first-line treatment for clinical malaria over 14 days is low (10%). Points in each panel show the median expected reduction in cumulative cases achieved by combining seasonal deployment of a pre-liver stage, blood stage, or multi-stage product with SMC, relative to cumulative cases when SMC is deployed alone. Median reductions are evaluated across all possible combinations of product initial efficacy, protection half-life, and decay shape. Bars indicate the 25% and 75% quantiles of the corresponding reductions.

**Figure 4.2:**
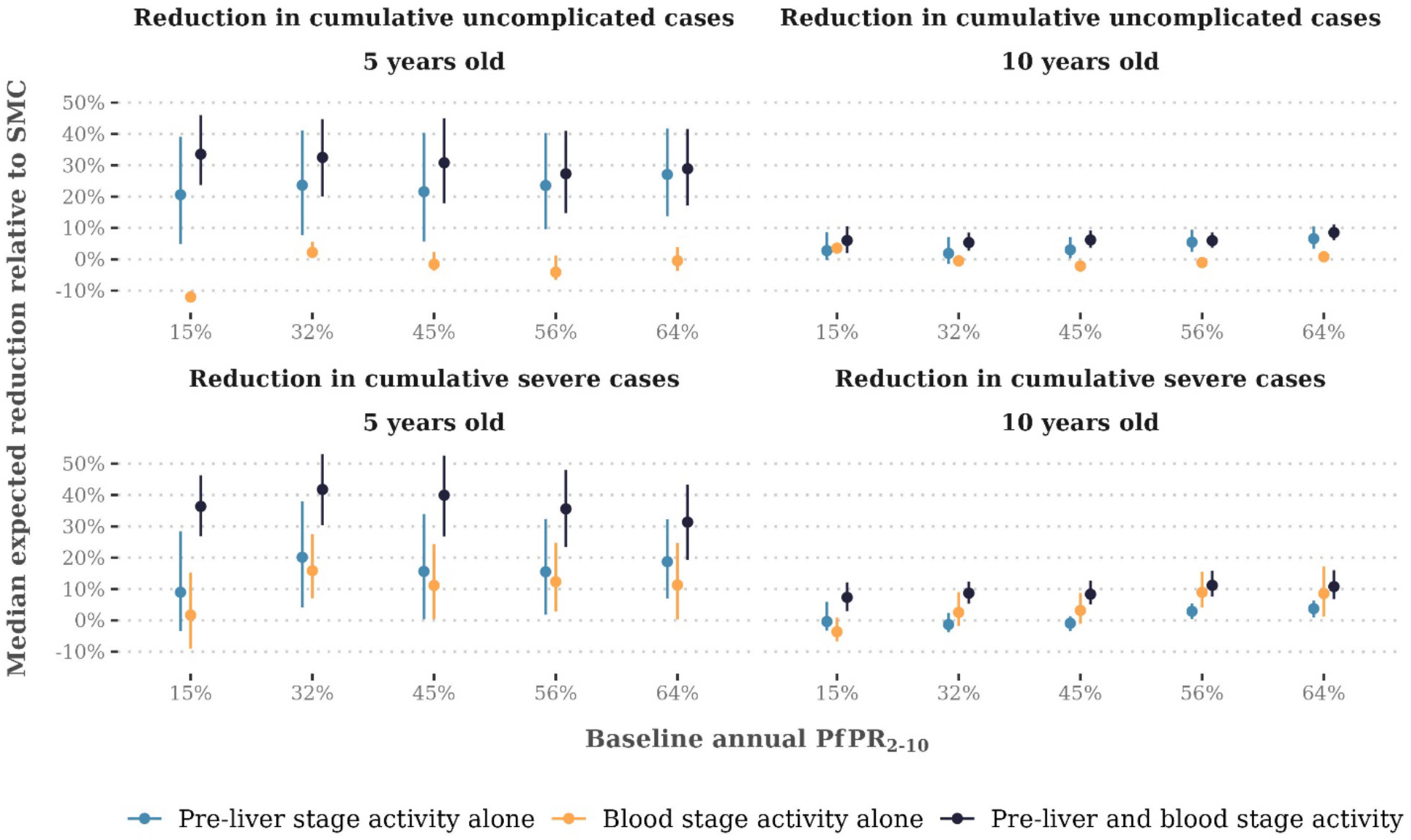
Differences in median impact on cumulative case outcomes for products with pre-liver stage activity alone, blood stage activity alone, and multi-stage pre-liver stage and blood stage activity, evaluated relative to perfect deployment of SMC

Results show the imperfect seasonal coverage scenario, where children aged three to 59 months received four SMC cycles. 75% of malaria cases occur within six months of the year and the probability of seeking first-line treatment for clinical malaria over 14 days is low (10%). Points in each panel show the median expected reduction in cumulative cases achieved by combining seasonal deployment of a pre-liver stage, blood stage, or multi-stage product with SMC, relative to cumulative cases when SMC is deployed alone. Median reductions are evaluated across all possible combinations of product initial efficacy, protection half-life, and decay shape. Bars indicate the 25% and 75% quantiles of the corresponding reductions.

**Figure 4.3:**
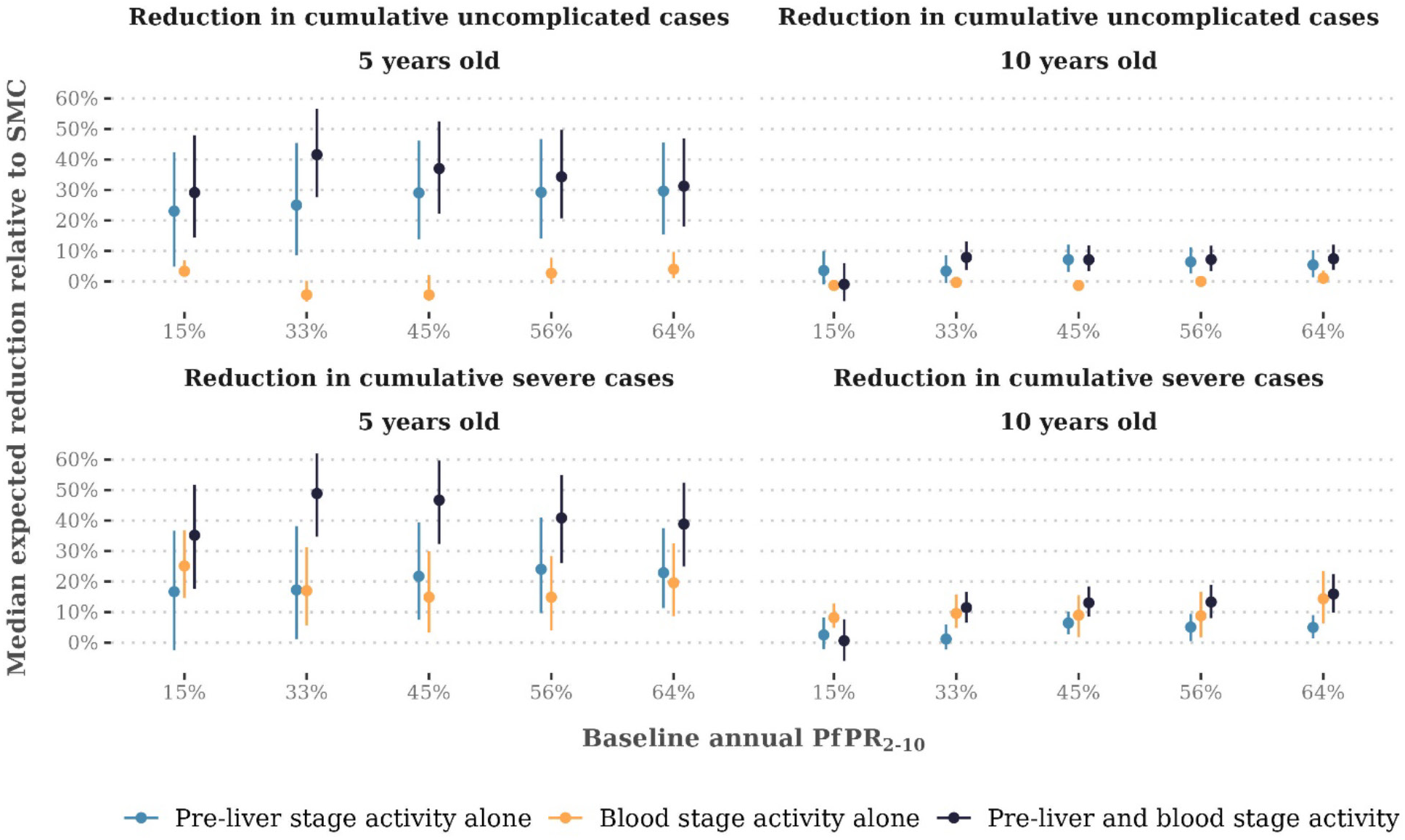
Differences in median impact on cumulative case outcomes for products with pre-liver stage activity alone, blood stage activity alone, and multi-stage pre-liver stage and blood stage activity, evaluated relative to imperfect seasonal coverage of SMC

Results show the random allocation scenario, where between 30% and 100% of children aged three to 59 months were randomly allocated to receive each of five SMC cycles. 75% of malaria cases occur within six months of the year and the probability of seeking first-line treatment for clinical malaria over 14 days is low (10%). Points in each panel show the median expected reduction in cumulative cases achieved by combining seasonal deployment of a pre-liver stage, blood stage, or multi-stage product with SMC, relative to cumulative cases when SMC is deployed alone. Median reductions are evaluated across all possible combinations of product initial efficacy, protection half-life, decay shape, and SMC cycle coverage. Bars indicate the 25% and 75% quantiles of the corresponding reductions.

**Figure 4.4:**
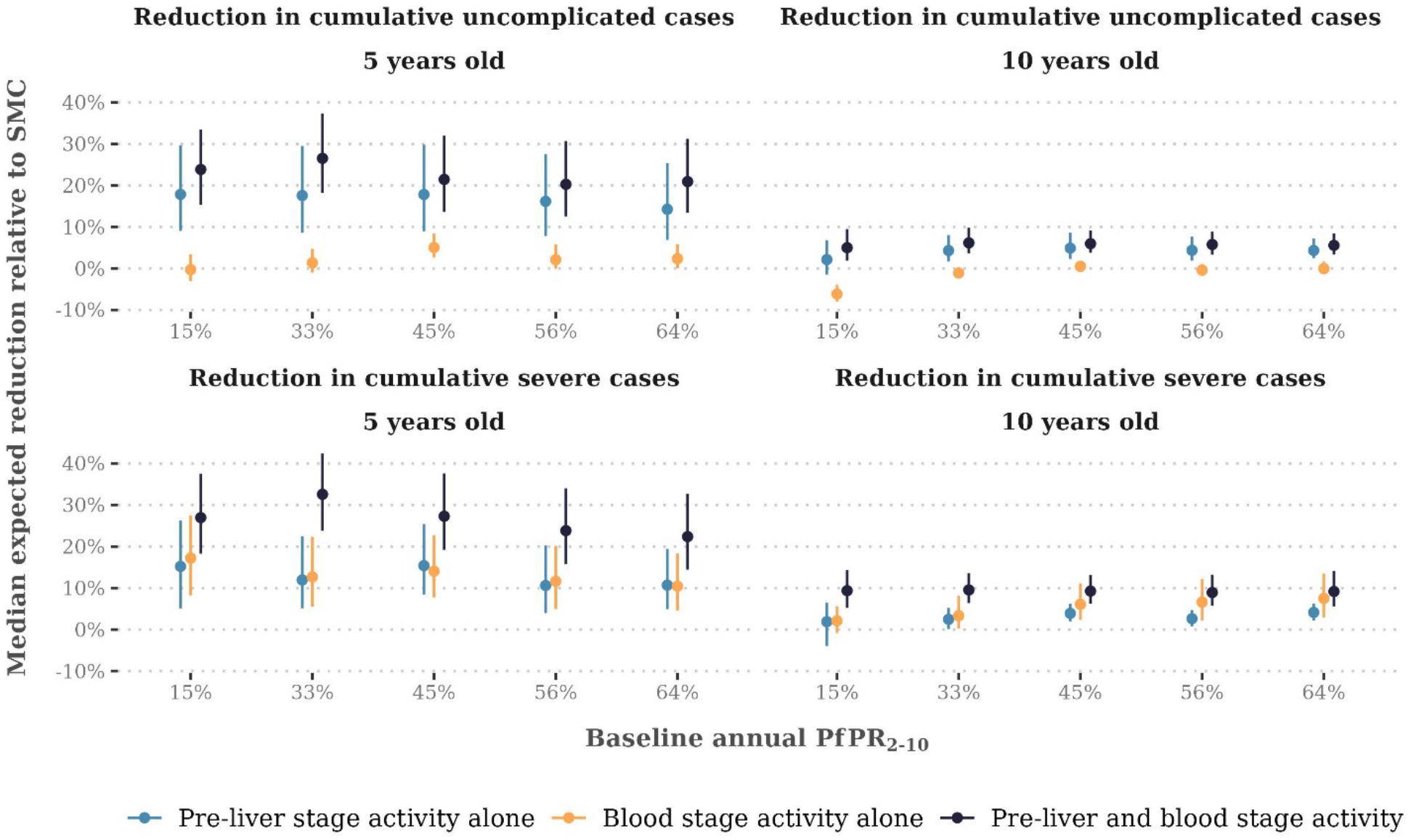
Differences in median impact on cumulative case outcomes for products with pre-liver stage activity alone, blood stage activity alone, and multi-stage pre-liver stage and blood stage activity, evaluated relative to random coverage of SMC cycles

